# Analysis of severe illness after post-vaccination COVID-19 breakthrough among adults with and without HIV in the United States

**DOI:** 10.1101/2022.04.15.22273913

**Authors:** Raynell Lang, Elizabeth Humes, Sally B. Coburn, Michael A. Horberg, Lily F Fathi, Eric Watson, Celeena R Jefferson, Lesley S. Park, Amy C. Justice, Kirsha S. Gordon, Kathleen M Akgün, Sonia Napravnik, Jessie K. Edwards, Lindsay E. Browne, Deana M. Agil, Michael J. Silverberg, Jacek Skarbinski, Wendy A. Leyden, Cameron Stewart, Brenna C Hogan, Kelly A. Gebo, Vincent C. Marconi, Carolyn F. Williams, Keri N. Althoff, the Corona-Infectious-Virus Epidemiology Team (CIVETs) of the NA-ACCORD of IeDEA

**Author notes:** **Corresponding author:** Keri N Althoff, PhD, MPH, Associate Professor, Department of Epidemiology, Johns Hopkins Bloomberg School of Public Health, 615 N Wolfe St, Rm E7142, Baltimore, MD 20295.

## Abstract

**Importance:** Understanding the severity of post-vaccination COVID-19 breakthrough illness among people with HIV (PWH) can inform vaccine guidelines and risk-reduction recommendations.

**Objective:** Estimate the rate and risk of severe breakthrough illness among vaccinated PWH and people without HIV (PWoH) who experience a breakthrough infection.

**Design, setting, and participants:** The Corona-Infectious-Virus Epidemiology Team (CIVET-II) collaboration consists of four US longitudinal cohorts from integrated health systems and academic centers. Adults (≥18 years old), in-care, fully vaccinated by June 30, 2021 with HIV, and matched PWoH (on date fully vaccinated, age group, race/ethnicity, and sex) were the source population. Those who experienced a post-vaccination SARS-CoV-2 breakthrough infection were eligible. Severe COVID-19 breakthrough illness was defined as hospitalization due to COVID-19. Discrete time proportional hazards models estimated adjusted hazard ratios (aHR) and 95% confidence intervals ([,]) of severe breakthrough illness by HIV status adjusting for demographics, COVID-19 vaccine type, and clinical factors. The proportion of patients requiring mechanical ventilation or died was compared by HIV status.

**Exposure:** HIV infection

**Outcome:** Severe COVID-19 breakthrough illness, defined as hospitalization within 28 days after a breakthrough SARS-CoV-2 infection with a primary or secondary COVID-19 discharge diagnosis.

**Results:** Among 1,241 PWH and 2,408 PWoH with breakthrough infections, the cumulative incidence of severe illness in the first 28 days was low and comparable between PWoH and PWH (7.3% vs. 6.7%, respectively, risk difference=-0.67% [-2.58%, 1.23%]). The risk of severe breakthrough illness was 59% higher in PWH with CD4 counts <350 cells/mm^3^ compared with PWoH (aHR=1.59 [0.99, 2.46]). In multivariable analyses among PWH, being female, older, having a cancer diagnosis, and lower CD4 count increased the risk of severe breakthrough illness, while previous COVID-19 reduced the risk. Among all patients, 10% were mechanically ventilated and 8% died, with no difference by HIV status.

**Conclusions and Relevance:** The risk of severe COVID-19 breakthrough illness within 28 days of a breakthrough infection was low among vaccinated PWH and PWoH. However, PWH with moderate and severe immune suppression had a higher risk of severe breakthrough infection. Recommendations for additional vaccine doses and risk-reduction strategies for PWH with moderate immune suppression may be warranted.

**Key Points:** *Question:* In 2021, among fully vaccinated people with COVID-19 breakthrough illness, was the risk of severe illness higher in people with HIV (PWH) compared to people without HIV (PWoH)?

*Findings:* PWH with <350 cells/mm^3^ have a 59% increased risk of severe breakthrough illness compared to PWoH.

*Meaning:* Vaccinations effectively reduce the risk of severe COVID-19 infection in both PWH and PWoH; however, PWH having a CD4 count <350 cells/mm^3^ are at higher risk of severe breakthrough infection compared to PWoH. PWH with moderate immune suppression should be considered for additional vaccine dosages and other risk-reduction measures.

## INTRODUCTION

SARS-CoV-2 vaccination is an effective protective measure against coronavirus disease 2019 (COVID-19).^1-3^ Although uncommon, people with HIV (PWH) have a higher risk for post-vaccination (i.e. breakthrough) SARS-CoV-2 infection.^4-9^ Our prior study showed a 28% increased risk for breakthrough SARS-CoV-2 infection in PWH compared to people without HIV (PWoH), although breakthrough cumulative incidence was low (PWH=3.1%, PWoH=2.5%), consistent with findings among people with other immunosuppressive conditions.^4,5,9^ Data on breakthrough COVID-19 illness, particularly severe illness requiring hospitalization, in PWH remain sparse^10,11^

Studies on the severity of breakthrough COVID-19 illness by HIV status are equivocal, some finding comparable severity,^12-15^ while others have reported a higher risk of developing severe illness and worse outcomes, including death, for PWH compared to PWoH.^16-19^ Immune dysfunction is believed to increase severe COVID-19 illness risk in PWH, with lower CD4 counts and detectable HIV viral loads associated with worse outcomes.^20,21^ A higher risk of severe illness in PWH may be confounded by the greater comorbidity burden among PWH (compared to similar aged PWoH), including hypertension, diabetes, cardiovascular disease, and smoking.^22,23^ Conversely, PWH have greater immune dysfunction and a lower likelihood of hyperactive cytokine response, which may reduce the risk of severe illness in PWH.^22^ With global COVID-19 vaccination uptake, there is an increasing need to estimate severe COVID-19 breakthrough illness rates and risk among PWH and an appropriate control group. However, large cohorts are needed to observe sufficient breakthrough infections that progress to severe illness.

Current US Centers for Disease Control and Prevention (CDC)’s guidelines recommend risk reduction behaviors (i.e. mask wearing), an additional COVID-19 primary series vaccine dose and second booster dose for PWH having “advanced or untreated HIV infection.”^24,25^ PWH having partially recovered CD4 counts and moderate immune suppression are not currently recommended for an additional or second booster dose. Our objective was to determine if HIV infection was associated with increased severe COVID-19 outcomes among fully vaccinated adults with a breakthrough SARS-CoV-2 infection, and to determine the risk factors for severe COVID-19 breakthrough illness among PWH.

## METHODS

### Study population

The Corona-Infectious-Virus Epidemiology Team (CIVET-II) cohort is comprised of four cohorts including Kaiser Permanente Mid-Atlantic States (Maryland, District of Columbia, northern Virginia), Kaiser Permanente Northern California, University of North Carolina Chapel Hill HIV Clinic (UNC), and the Veterans Aging Cohort Study (VACS), a cohort of PWH (and similar PWoH) receiving care within the National US Veterans Affairs Healthcare System. This collaboration is an extension of the North American AIDS Cohort Collaboration on Research and Design.^26^ Each local institution and the Johns Hopkins Bloomberg School of Public Health institutional review board granted approval.

Adults (≥18 years old), “in-care” (**Supplement Table 1**), and fully vaccinated against COVID-19 between December 11, 2020 (Emergency Use Authorization of the first COVID-19 vaccine) and June 30, 2021, were eligible. Full vaccination status was defined using CDC criteria: a) 14 days after BNT162 (Pfizer) or mRNA-1273 (Moderna) mRNA vaccine second dose; or b) 14 days after Janssen Ad26.COV2.S (J&J) single dose.^27^ Patients with vaccines not authorized in the US were excluded.

Fully vaccinated PWH were matched to three PWoH on the date fully vaccinated (+/-14 days of the PWH vaccination date), 10-year age group, race/ethnicity, and sex at birth. Race/ethnicity was included given the disproportionate burdens of HIV by race/ethnicity. To maximize the cohort size, PWH could be matched to individuals one age group above or below their category. If three matches were unavailable, PWH were matched to one or two PWoH. The VACS (N=67,627) matches each Veteran with HIV to two Veterans without HIV per their longstanding schema, based on age, race/ethnicity, sex, and clinical site at cohort entry; VACS participants were not matched on date fully vaccinated.^28^

Participants were eligible for the present study if they had breakthrough COVID-19, defined as the first SARS-CoV-2 infection (detectable SARS-CoV-2 nucleic acid amplification assay [NAAT] or antigen test) or COVID-19 diagnosis (International Classification of Diseases (ICD)-10 codes [**Supplement Table 2**]) after the date fully vaccinated. Additional detectable SARS-CoV-2 laboratory test results and/or diagnoses occurring within +/- 90 days of breakthrough were considered persistent infection.^29^ If a COVID-19 diagnosis code was also identified within the 90-day window of detectable result, the first laboratory test was the breakthrough diagnosis date.

All variables were abstracted from electronic health records (EHR). Our study follows the Strengthening the Reporting of Observational Studies in Epidemiology (STROBE) reporting guidelines.

### Outcome: Severe COVID-19 breakthrough illness

Severe COVID-19 breakthrough illness was defined as: 1) hospitalization within 28 days of breakthrough; and 2) discharge diagnosis ranked first or second was COVID-19. All discharge diagnoses were evaluated by two infectious disease physicians to exclude hospitalizations that were unlikely due to COVID-19 (trauma, surgical, non-COVID-19 infections, mental health, or substance use admissions). Only the first-ranked discharge diagnosis was available for one cohort (contributing 28% of the study population). In this cohort, if COVID-19 was not the first-ranked discharge diagnosis, and there was ≥1 other discharge diagnosis suggestive of COVID-19 (i.e., pneumonia due to coronavirus disease, other viral pneumonia, acute respiratory failure, or hypoxemia) the patient was classified as the outcome of severe COVID-19 breakthrough illness.

Mechanical ventilation and extracorporeal membrane oxygenation (ECMO) procedures were extracted from ICD-10 procedure and current procedural terminology (CPT) codes (**Supplement Table 2**) occurring between the hospital admission and discharge dates. Death occurring during or within 30 days following hospitalization or within 30 days following COVID-19 diagnosis among those not hospitalized was extracted from EHR.

### Exposure: HIV infection

PWH were identified using HIV registries or HIV ICD diagnosis codes (**Supplement Table 1**). PWoH were classified as such if there was no evidence of HIV infection using these same sources as of December 11, 2020.

### Covariates

Demographic covariates included age, race/ethnicity, and sex at birth. COVID-19 covariates included the primary series vaccine type (Pfizer, Moderna, J&J), additional vaccine dose (receipt ≥28 days after completion of primary series), and SARS-CoV-2 infection prior to date fully vaccinated (history of COVID-19). History of COVID-19 included both infections occurring prior to any vaccination or in the window between the first dose and full vaccination (partial breakthrough).

Comorbidity covariates included obesity (body mass index [BMI] ≥30 kg/m^2^), type 2 diabetes (DM), hypertension (HTN), end-stage renal disease (ESRD), immune suppressive conditions (organ or tissue transplantation [SOT], rheumatoid arthritis [RA], systemic lupus erythematosus [SLE]), cancer, and pregnancy. Comorbidity diagnoses were identified using ICD-10 codes, measured closest to the date fully vaccinated and after October 1, 2015; or January 1, 2020, for cancer and pregnancy only (see **Supplement Table 2** for ICD-10 diagnosis codes).

Among PWH, CD4 T-cell count, and HIV-1 plasma RNA viral load suppression were collected closest to the date of full vaccination after January 1, 2020, and at antiretroviral therapy initiation (12 months prior to 1 month after). HIV viral suppression was defined as <50 copies/mL. History of AIDS diagnosis (clinical diagnosis^30^ or CD4 count <200 cells/mm^3^) prior to date fully vaccinated was included.

### Statistical analysis

Study entry was the date of observed breakthrough COVID-19. Individuals were followed to date of severe breakthrough COVID-19 illness (outcome) or date of death, disenrollment from the health system (applicable to 2 of the health systems), 28 days after breakthrough COVID-19, or December 31, 2021, whichever occurred first.

Severe COVID-19 breakthrough illness monthly incidence rates (IR) per 100 person-years (PY) and 95% confidence intervals ([,]) were calculated by HIV status. Severe COVID-19 breakthrough illness cumulative incidence was estimated from the date of breakthrough COVID-19 through day 28; estimates were stratified by HIV status, and among PWH, CD4 count (<350, 350-499, and ≥500 cells/mm^3^) and viral suppression status. Using the same timescale, cumulative incidence was also estimated by HIV status for each vaccine type and for those who received an additional vaccine dose (≥28 days after primary series completion and prior to breakthrough COVID-19). Log-rank tests were calculated to test for differences in cumulative incidence and risk differences were estimated (with standard error or Wald 95% confidence intervals).

A discrete time-to-event analysis using a complimentary log-log model estimated the unadjusted and adjusted hazard ratios (aHR) with 95% confidence intervals for severe COVID-19 breakthrough illness risk by HIV status. Adjustment factors included: sex, race/ethnicity, age, additional vaccine dose following primary series, prior COVID-19, obesity, DM, HTN, ESRD, SOT, RA, SLE, cancer, and cohort. Subgroup analyses stratified by HIV status allowed for investigations into risk factors for severe COVID-19 breakthrough illness in both groups. Among PWH, prior AIDS diagnosis, HIV viral suppression and CD4 count were evaluated as risk factors for severe COVID-19. Sensitivity analyses excluded participants without first or second diagnosis code rankings.

## RESULTS

Among 113,994 patients (33,029 PWH and 80,965 PWoH), there were 3,649 breakthrough COVID-19 infections (1,241 PWH and 2,408 PWoH). Of those who experienced a COVID-19 breakthrough infection, 60% were >55 years, 89% were male, 47% were non-Hispanic Black, 59% received Pfizer and 31% received Moderna for their primary series, and 15% received an additional COVID-19 vaccine dose ≥28 days after primary series completion (20% in PWH and 13% in PWoH). PWH who had CD4 count of <200 cells/mm^3^ (17%) and 200-349 cells/mm^3^ (16%) were less likely to receive an additional vaccine dose, compared to PWH with CD4 counts ≥350 cells/mm^3^ (20%) (**Supplemental Table 3**). Among the N=3,649 patients with breakthrough COVID, the majority (75%) were laboratory confirmed and the remainder were based on clinical diagnosis codes. Forty-nine percent of breakthroughs occurred during the Delta variant (B.1.617.2) surge from July-Oct 2021, and 41% occurred from Nov-Dec 2021 (the start of the Omicron variant [B.1.1.529] wave) (**Table 1**). PWH (compared with PWoH) had a lower proportion with obesity (36% vs. 54%), DM (25% vs. 34%), and HTN (51% vs. 60%) prior to vaccination. The proportion with ESRD, RA, SLE, or having received a SOT or a cancer diagnosis were similar by HIV status. Among PWH, 26% had history of AIDS prior to vaccination, 91% were virally suppressed and median CD4 count was 620 (interquartile range [IQR]: 438, 846) cells/mm^3^ at the time fully vaccinated.

**Table 1:** Characteristics at date of SARS-CoV-2 breakthrough infection, N=3,649

| Characteristic | PWoH, N = 2,408<sup>1</sup> | PWH, N = 1,241<sup>1</sup> |
| --- | --- | --- |
| **Hospitalization** | 170 (7%) | 79 (6%) |
| **Mechanical Ventilation among those hospitalized** | 16 (9%) | 8 (10%) |
| **Death among breakthrough infections** | 21 (1%) | 12 (1%) |
| **Age** |  |  |
| 18-24 | 9 (0%) | 9 (1%) |
| 25-34 | 137 (6%) | 112 (9%) |
| 35-44 | 279 (12%) | 184 (15%) |
| 45-54 | 492 (20%) | 245 (20%) |
| 55-64 | 797 (33%) | 359 (29%) |
| 65-74 | 539 (22%) | 264 (21%) |
| 75+ | 155 (6%) | 68 (6%) |
| **Sex** |  |  |
| Male | 2,108 (88%) | 1,136 (92%) |
| Female | 300 (13%) | 105 (9%) |
| **Ethnicity and Race** |  |  |
| Non-Hispanic white | 744 (31%) | 421 (34%) |
| Non-Hispanic Black/African American | 1,151 (48%) | 555 (45%) |
| Hispanic | 366 (15%) | 183 (15%) |
| Other | 131 (5%) | 71 (6%) |
| Unknown | 16 (1%) | 11 (1%) |
| **Month of breakthrough** |  |  |
| Jan-June | 247 (10%) | 134 (11%) |
| July-Oct | 1,132 (47%) | 639 (51%) |
| Nov-Dec | 1,029 (43%) | 468 (38%) |
| **Primary vaccination series type** |  |  |
| Pfizer | 1,194 (50%) | 605 (49%) |
| Pfizer + 3rd dose | 194 (8%) | 165 (13%) |
| Moderna | 676 (28%) | 301 (24%) |
| Moderna + 3rd dose | 95 (4%) | 68 (6%) |
| J&J | 234 (10%) | 93 (7%) |
| J&J + 2nd dose | 15 (1%) | 9 (1%) |
| **COVID prior to fully vaccinated** | 292 (12%) | 176 (14%) |
| **Comorbidities at fully vaccinated** |  |  |
| Obese BMI ($\geq 30$ kg/m<sup>2</sup>) | 1,218 (54%) | 434 (36%) |
| Diabetes | 817 (34%) | 308 (25%) |
| Hypertension | 1,443 (60%) | 633 (51%) |
| End-stage renal disease | 63 (3%) | 38 (3%) |
| **Immune suppressive conditions at fully vaccinated** |  |  |
| Organ or tissue transplantation | 43 (2%) | 18 (2%) |
| Rheumatoid arthritis | 44 (2%) | 14 (1%) |
| Lupus | 10 (0%) | 3 (0%) |
| Cancer diagnosis | 187 (8%) | 129 (10%) |
| Pregnancy | 4 (0%) | 3 (0%) |
| **CD4 at ART initiation (cells/mm<sup>3</sup>)** |  | 393(234, 613) |
| unknown |  | 536 (43%) |
| **AIDS before fully vaccinated** |  | 325 (26%) |
| **CD4 at fully vaccinated (cells/mm<sup>3</sup>)** |  | 620 (438, 846) |
| <200 |  | 54 (4%) |
| 200-349 |  | 109 (9%) |
| 350-499 |  | 182 (15%) |
| ≥500 |  | 707 (57%) |
| unknown |  | 189 (15%) |
| **Suppressed HIV RNA at fully vaccinated (<50 copies/mL)** |  | 1,016 (91%) |
| unknown |  | 120 (10%) |

<sup>1</sup>n (%) except for CD4 counts where the medians (interquartile ranges) are reported
Definitions:
- Death defined as died while hospitalized for COVID-19 or within 30 days following discharge or within 30 days of COVID-19 diagnosis among those not hospitalized. - Mechanical Ventilation: Measured during the dates of hospital admission and discharge. - Age was categorized in 5-year increments for descriptive purposes - BMI, Diabetes, Hypertension, ESRD, Organ or tissue transplantation, RA, Lupus: Measured as close to the date fully vaccinated as possible within the window of 10/1/2015 – date fully vaccinated. - Cancer, Pregnancy: Measured as close to the date fully vaccinated as possible within the window of 1/1/2020 – date fully vaccinated - AIDS was defined using clinical diagnosis codes of AIDS-defining conditions.
AIDS, acquired immunodeficiency syndrome ART, antiretroviral Therapy BMI, body mass index COVID-19, Coronavirus Disease 2019 ESRD, end-stage renal disease HIV, human immunodeficiency virus PWH, people with HIV PWoH, people without HIV RA, rheumatoid arthritis

Among those with breakthrough COVID-19, 249 (7% overall; 79 or 6% of PWH; 170 or 7% PWoH) had severe COVID-19 breakthrough illness and were hospitalized (**Table 1** and **Supplement Figure 1**). Most (66%) hospitalizations occurred on the same day, and 81% occurred within two days following COVID-19 diagnosis. The median duration of hospitalization was 5 (IQR: 3, 8) days among PWoH and 4 (IQR: 2, 8) days among PWH. Among all hospitalized patients 10% were mechanically ventilated (**Table 1**). There were 33 deaths (1% overall; 12 or 1% in PWH; 21 or 1% in PWoH) that occurred during or within 30 days of hospital discharge or COVID-19 diagnosis. **Supplement Table 4** describes characteristics stratified by HIV status and severe COVID-19 breakthrough illness status.

### Incidence Rates (IR) and Cumulative Incidence of Severe COVID-19 Breakthrough Illness

The IR of severe COVID-19 breakthrough illness was higher among PWoH (138 [118, 160] /100 PY) versus PWH (117 [92, 145] /100 PY) (**Figure 1**) and relatively stable over time (**Figure 1, Supplement Table 5**) with slight fluctuations reflecting the bimodal distribution of the Delta and Omicron variant waves (**Supplement Figure 2**).

**Figure 1:**
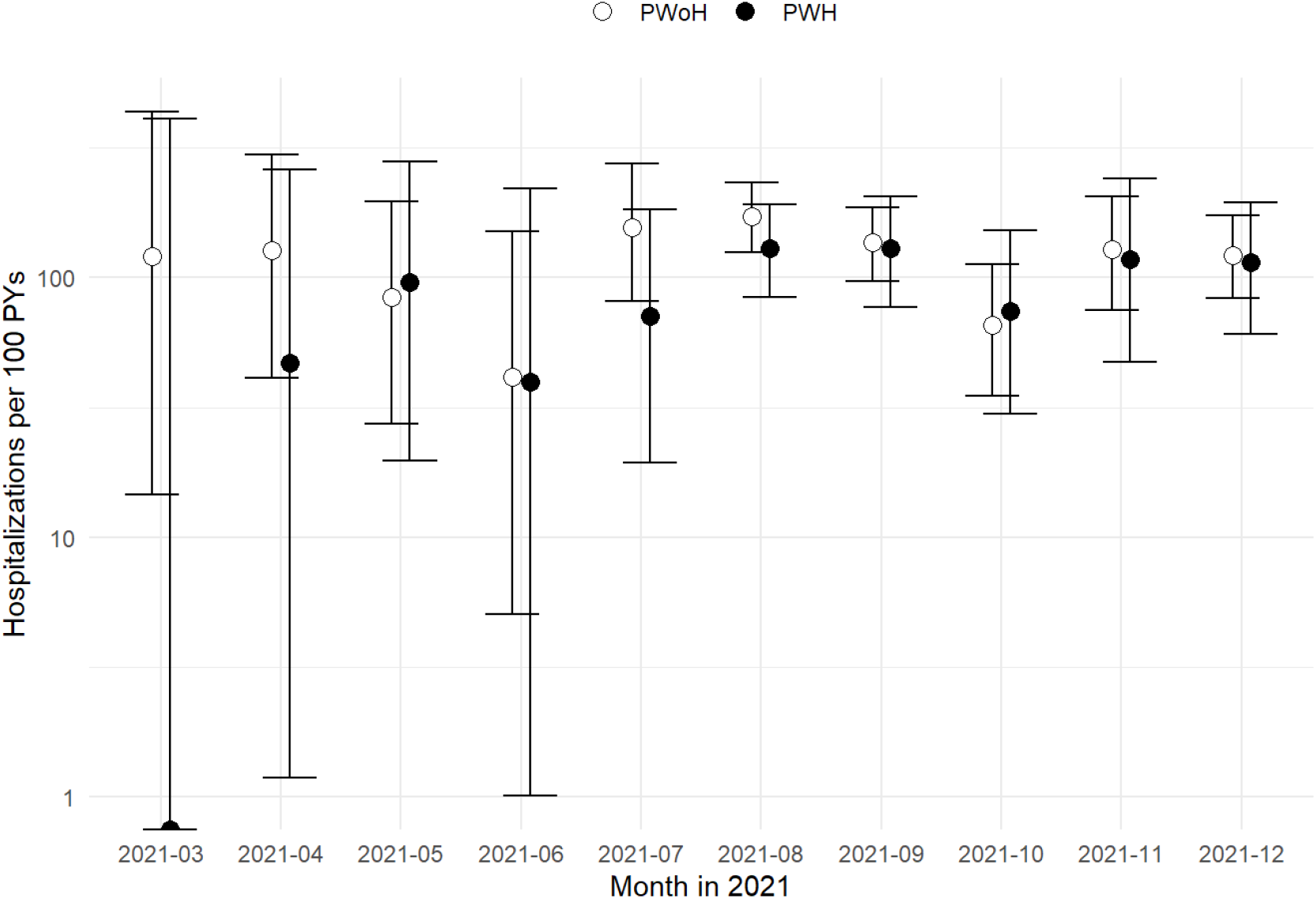
Incidence Rates of Severe COVID-19 breakthrough per 100 patient-years (PY) in people with and without HIV by month (along with 95% CI) (N=3,649) Footnote: The incidence rate estimates for January 2021 were not estimated as there were 0 and 0.02 person-years of observation after COVID-19 breakthrough infection in PWH and PWoH, respectively. Similarly, the person years in February 2021 were 0.14 and 0.21 in PWH and PWoH (respectively) and there were no severe COVID-19 breakthrough illness events; incidence rates were not estimated.

The 28-day severe COVID-19 breakthrough illness cumulative incidence was similar among PWoH (7.3% [6.3%, 8.4%]) versus PWH (6.7% [5.2% 8.1%], log rank p=0.399, risk difference=-0.67% [-2.58%, 1.23%]) (**Figure 2a**). PWH with lower CD4 counts (<350 cells/mm^3^) at full vaccination had a higher risk of severe COVID-19 breakthrough illness compared with both PWoH and PWH with CD4 counts vs. >350 cells/mm^3^ (**Figure 2b, Supplement Figure 3**). Risk did not differ by HIV viral load (Figure 2c).

**Figure 2:**
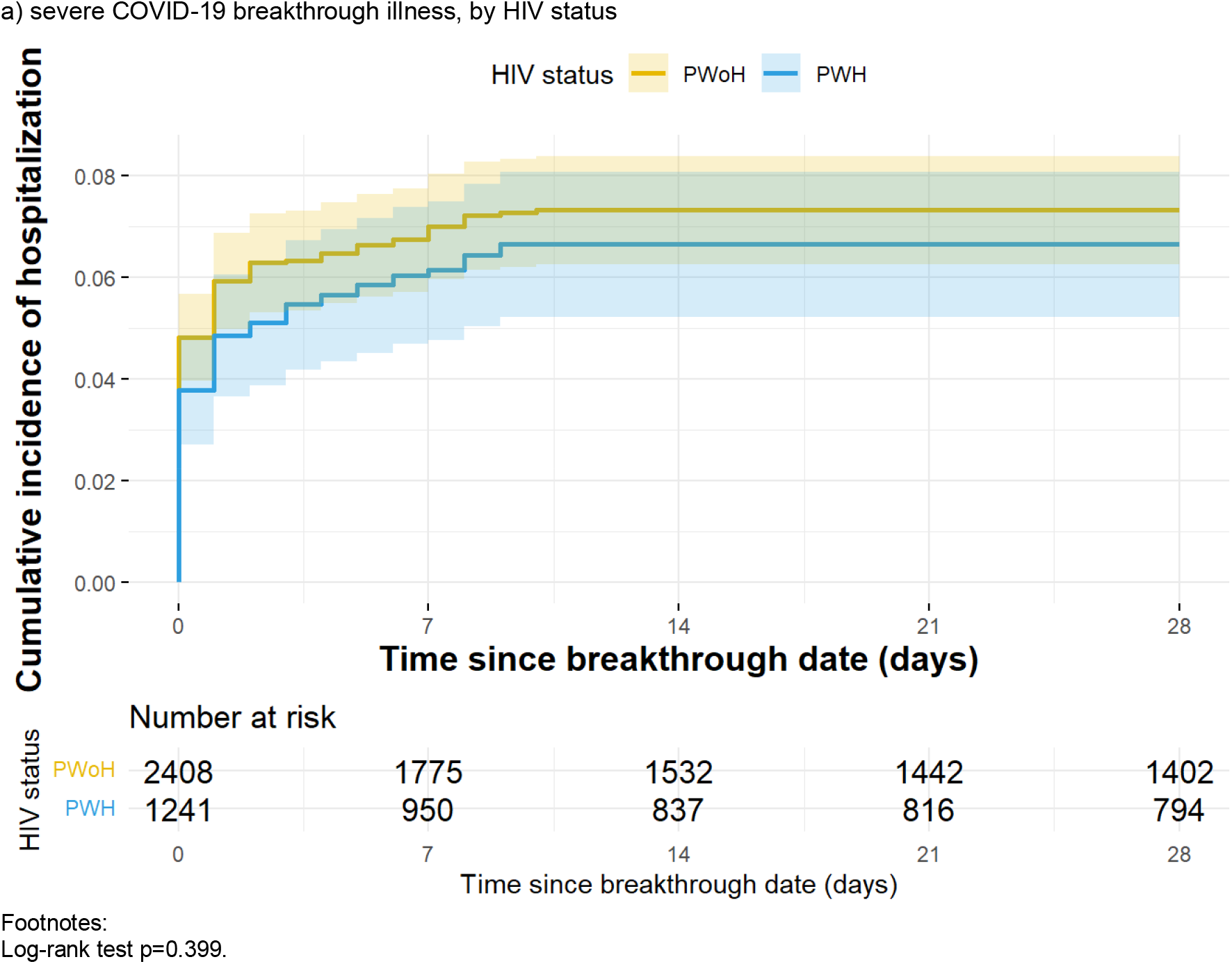
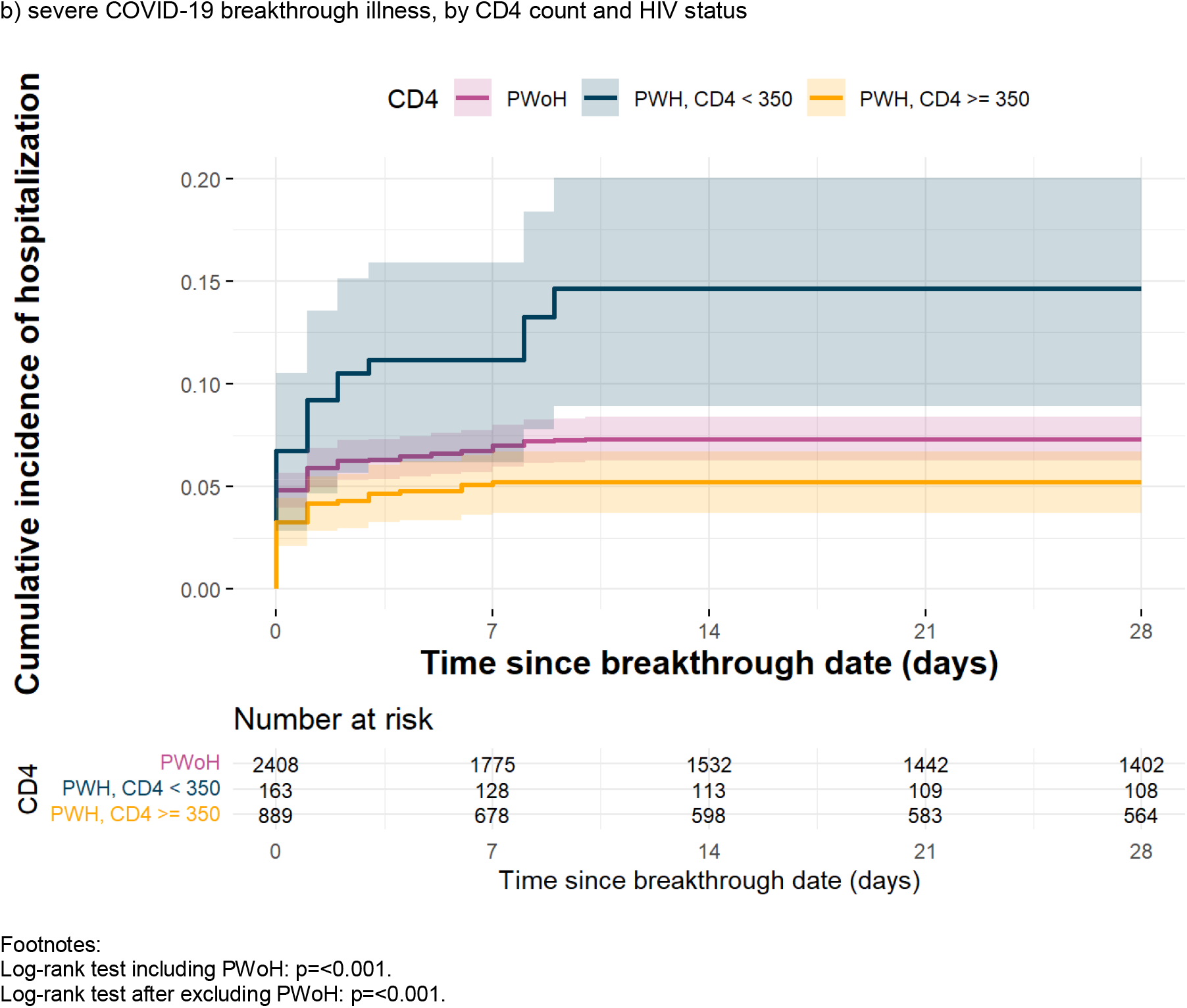
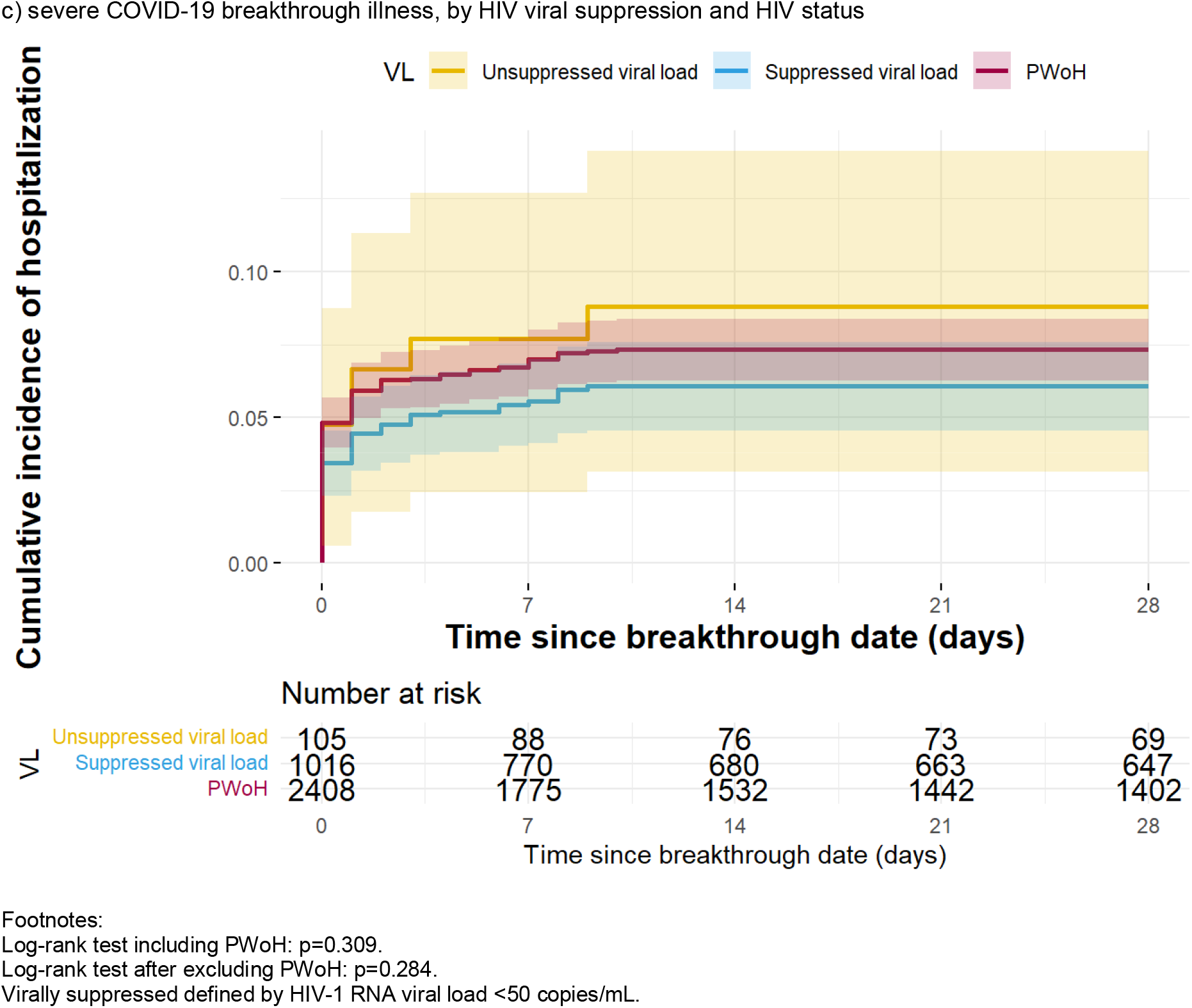
Cumulative incidence of severe COVID-19 breakthrough illness (and 95% confidence intervals represented by the shading), stratified by a) HIV status, b) CD4 count and HIV status, and c) HIV viral suppression and HIV status

Severe COVID-19 breakthrough illness risk was highest among patients with J&J for their primary vaccine series (9.3% [6.1, 12.4%]), followed by Pfizer (7.2% [6.1%, 8.3%]) and Moderna (6.2% [4.8%, 7.7%]) (**Figure 3a**), with no statistically significant differences by HIV status within each vaccine group. Regardless of the primary vaccine series type, having an additional dose reduced the risk of severe COVID-19 breakthrough illness in both PWH and PWoH (log-rank p=0.021, **Figure 3b**)

**Figure 3:**
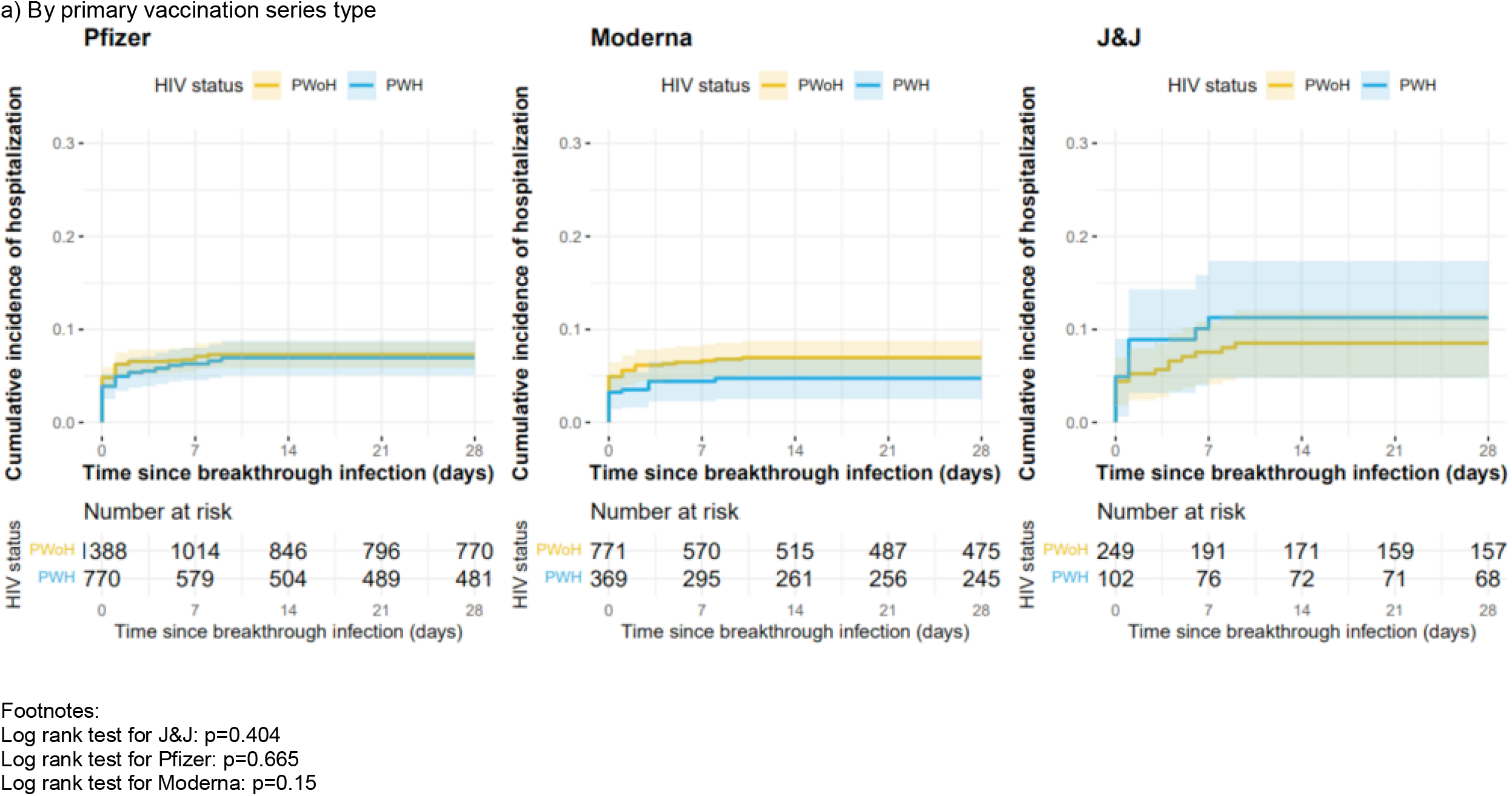
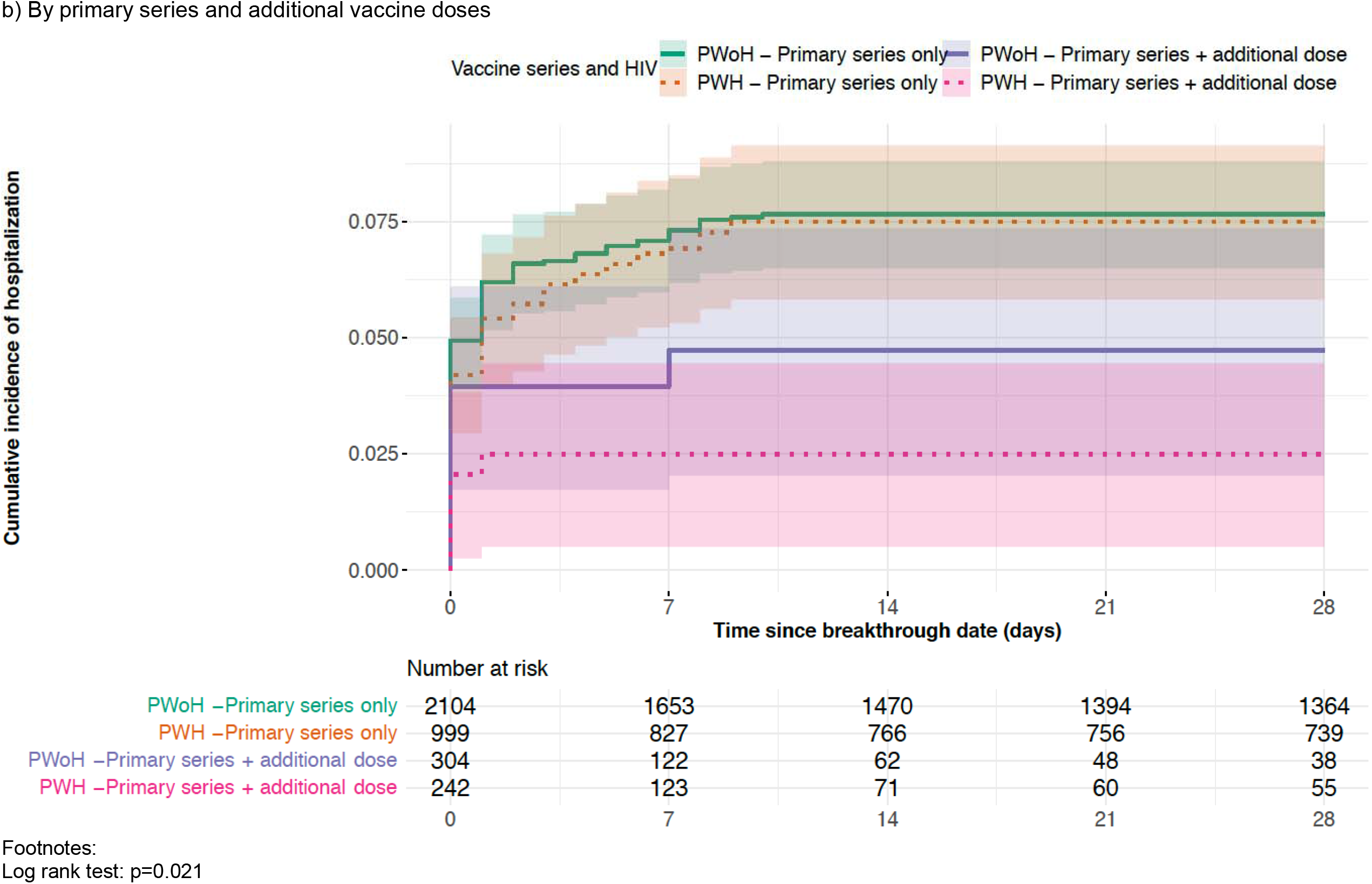
Cumulative incidence of severe COVID-19 breakthrough illness (and 95% confidence intervals represented by the shading), stratified by HIV status and primary vaccination series type

### Risk factors for Severe COVID-19 Breakthrough Illness, by HIV status

There was no difference in severe COVID-19 breakthrough illness risk in PWH vs. PWoH (aHR=1.02 [0.76, 1.35]); however, there was a 59% increased risk among PWH having a CD4 count <350 cells/mm^3^ compared with PWoH (aHR=1.59 [0.99, 2.46], **Table 2**). Stratified by HIV status, PWH and PWoH had an increased risk of severe COVID-19 breakthrough illness with increasing age and decreased risk among those with an additional vaccine dose.

**Table 2:** Crude (HR) and adjusted hazard ratios (aHR) and 95% confidence intervals (CI) of severe SARS-CoV-2 breakthrough illness

| HIV status | Total Population (N=3,385) |  |  |  |
| --- | --- | --- | --- | --- |
|  | HR (95% CI) |  | <sup>a</sup>aHR (95% CI) |  |
| PWoH | 1.00 |  | 1.00 |  |
| PWH, CD4 count <350cells/mm<sup>3</sup> | **2.00 (1.26, 3.02)** |  | **1.59 (0.99, 2.46)** |  |
| PWH, CD4 count ≥350cells/mm<sup>3</sup> | **0.70 (0.50, 0.97)** |  | 0.95 (0.67, 1.32) |  |
| Characteristics | <sup>a</sup> PWoH (N=2,350 n=170) |  | <sup>b</sup> PWH (N=1,035 n=68) |  |
|  | HR (95% CI) | aHR (95% CI) | HR (95% CI) | aHR (95% CI) |
| **Sex** |  |  |  |  |
| Male | 1.00 | 1.00 | 1.00 | 1.00 |
| Female | **0.25 (0.10, 0.52)** | 0.48 (0.16, 1.12) | 1.50 (0.66, 2.95) | **2.88 (1.03, 7.11)** |
| **Ethnicity and Race** |  |  |  |  |
| Non-Hispanic white |  | 1.00 | 1.00 | 1.00 |
| Non-Hispanic Black/African American | 1.24 (0.88, 1.77) | 1.26 (0.86, 1.85) | 1.26 (0.73, 2.22) | 1.26 (0.70, 2.32) |
| Hispanic | 1.01 (0.61, 1.62) | 1.21 (0.72, 1.98) | 0.98 (0.43, 2.10) | 1.54 (0.65, 3.38) |
| Other/Unknown | 0.52 (0.18, 1.19) | 1.24 (0.43, 2.86) | 1.30 (0.43, 3.21) | 2.71 (0.87, 7.12) |
| **Age** |  |  |  |  |
| <55 | 1.00 | 1.00 | 1.00 | 1.00 |
| 55-64 | **6.63 (3.54, 13.80)** | **3.49 (1.74, 7.86)** | **3.02 (1.52, 6.33)** | 2.11 (0.98, 4.80) |
| 65-74 | **12.60 (6.82, 26.00)** | **5.37 (2.64, 12.20)** | **4.41 (2.25, 9.13)** | **3.01 (1.37, 6.98)** |
| 75+ | **21.00 (10.70, 45.00)** | **8.44 (3.90, 20.00)** | **7.71 (3.25, 17.9)** | **5.21 (1.88, 14.40)** |
| **Primary vaccination series type** |  |  |  |  |
| Primary Series only | 1.00 | 1.00 | 1.00 | 1.00 |
| Primary Series + additional dose | 0.63 (0.34, 1.07) | **0.56 (0.29, 0.98)** | **0.43 (0.17, 0.91)** | 0.47 (0.18, 1.04) |
| **COVID-19 prior to fully vaccinated** | **0.39 (0.18, 0.71)** | **0.21 (0.09, 0.40)** | **0.43 (0.15, 0.97)** | **0.22 (0.07, 0.52)** |
| **Calendar period of breakthrough** |  |  |  |  |
| Jan – June | 1.00 |  | 1.00 |  |
| July – Oct | 1.49 (0.91, 2.62) |  | **2.53 (1.03, 8.40)** |  |
| Nov – Dec | 0.77 (0.45, 1.40) |  | 1.41 (0.53, 4.89) |  |
| **Comorbidities before fully vaccinated** |  |  |  |  |
| Obese BMI (≥30 kg/m<sup>2</sup> vs. <30) | 0.98 (0.72, 1.34) | 1.01 (0.73, 1.40) | 1.38 (0.85, 2.23) | 1.46 (0.85, 2.49) |
| Diabetes (vs. no) | **2.80 (2.07, 3.81)** | 1.11 (0.79, 1.56) | **2.26 (1.39, 3.64)** | 1.47 (0.85, 2.50) |
| Hypertension (vs. no) | **7.73 (4.65, 14.00)** | **4.51 (2.35, 9.80)** | **3.02 (1.78, 5.39)** | 1.27 (0.67, 2.50) |
| End-stage renal disease (vs. no) | **6.25 (3.88, 9.58)** | **2.53 (1.33, 4.56)** | 1.82 (0.55, 4.41) | 1.12 (0.31, 3.09) |
| **Immune suppressive conditions at fully vaccinated (vs. no)** |  |  |  |  |
| Organ or tissue transplantation | **6.79 (3.89, 11.0)** | **2.49 (1.20, 4.96)** | 0.95 (0.05, 4.27) | 0.55 (0.03, 3.39) |
| Rheumatoid arthritis | 2.03 (0.80, 4.19) | 1.33 (0.50, 2.87) | 1.28 (0.07, 5.78) | 1.09 (0.06, 5.45) |
| Cancer diagnosis | **2.64 (1.75, 3.86)** | 1.47 (0.95, 2.19) | **2.92 (1.64, 4.95)** | **1.97 (1.05, 3.51)** |
| Lupus<sup>c</sup> | 2.86 (0.47, 8.95) |  |  |  |
| **AIDS diagnosis before fully vaccinated** |  |  | **1.83 (1.11, 2.96)** | 1.22 (0.70, 2.10) |
| **HIV RNA at fully vaccinated** |  |  |  |  |
| Suppressed (<50 copies/mL) |  |  | 1.00 |  |
| Unsuppressed (≥50 copies/mL) |  |  | 1.43 (0.66, 2.74) |  |
| **CD4 count at fully vaccinated (cells/mm<sup>3</sup>)** |  |  |  |  |
| ≥500 |  |  | 1.00 | 1.00 |
| 350-499 |  |  | 1.28 (0.62, 2.44) | 1.19 (0.56, 2.35) |
| 200-349 |  |  | **2.52 (1.28, 4.66)** | 1.65 (0.80, 3.21) |
| <200 |  |  | **3.98 (1.86, 7.76)** | **2.57 (1.15, 5.29)** |

Total number of severe COVID-19 illness events=238.
Abbreviations: HR=crude hazard ratio. aHR=adjusted hazard ratio. 95% CI=95% confidence interval.
<sup>a</sup>Adjusted for age (categorized into a 4-level group to assess a dose response relationship among older people (i.e., 55+) that are known to be at greater risk), sex, race and ethnicity, primary vaccination series type, COVID-19 prior to fully vaccinated, Comorbidities before fully vaccinated, immune suppressive conditions at fully vaccinated, and cohort.
<sup>b</sup> Adjusted for the covariates in the table and cohort. 184 (15.1% of all PWH) were excluded due to missing CD4 or HIV RNA measurements.
<sup>c</sup> There were no severe COVID-19 breakthrough illnesses among PWH with lupus, and it was therefore excluded from the multivariable model.
Values with P<0.05 are bolded.

Among PWH, severe COVID-19 breakthrough illness risk increased with decreasing CD4 count (compared to CD4 ≥500 cells/mm^3^: 200-349 cells/mm^3^ aHR=1.65 [0.80, 3.21]; <200 cells/mm^3^ aHR=2.57 [1.15, 5.29]; **Table 2**). Female PWH had a nearly 3-fold increased severe COVID-19 breakthrough risk compared to males. Increased risk associated with non-Hispanic Black and Hispanic race/ethnicity and comorbidities ranged from 12% to 54% (p>0.05). Having a cancer diagnosis was associated with nearly 2-fold increased risk of severe COVID-19 breakthrough illness (aHR=1.97 [1.05, 3.51]).

Among PWoH, females had a reduced risk of severe COVID-19 breakthrough illness compared with males (p>0.05, **Table 2**). There was no observed difference in risk by race, obesity, DM, RA, SLE or having a cancer diagnosis; but there was a 4.5-fold (aHR=4.51 [2.35, 9.80]) increased risk with HTN and 2.5-fold increased risk with ESRD (aHR=2.53 [1.33, 4.56]) and SOT (aHR=2.49 [1.20, 4.96]).

Sensitivity analyses excluding patients without diagnosis code rankings did not qualitatively change findings.

### Mechanical ventilation, and death in severe COVID-19 breakthrough illness

Among the 249 hospitalized patients, a greater proportion of patients with CD4 counts <350 cells/mm^3^ required mechanical ventilation or died during hospitalization compared to PWH with higher CD4 counts and PWoH (Supplement **Figure 4**). No patients received ECMO.

Patients who needed mechanical ventilation (n=24) or died (n=33) were older (≥55 years), male, non-Hispanic Black, had high proportions of comorbidities, and low uptake (10-13%) of additional vaccine doses **(Supplemental Table 6)**. Of those who had a known death during or within 30 days following COVID-19 hospitalization (n=33), the majority had comorbidities including obese BMI (68%), HTN (95%), DM (65%), ESRD (30%), SOT (20%), RA (10%), or a cancer diagnosis (25%). Among PWH who died during a severe COVID-19 breakthrough illness (n=12), 50% had a prior diagnosis of AIDS and their median CD4 count at fully vaccinated was 352 (IQR 291, 423) cells/mm^3^.

## DISCUSSION

Prior CIVETs collaboration analyses showed a 28% increase in breakthrough COVID-19 among PWH compared with PWoH.^4^ Our present findings show the risk of severe illness (requiring hospitalization) after COVID-19 breakthrough was low (7% of 3,649 vaccinated PWH and PWoH) and did not differ by HIV status overall. PWH with lower CD4 counts (<350cells/mm^3^), however, had a 59% increase in the risk of severe COVID-19 breakthrough illness compared to PWoH, suggesting a role of immune dysfunction in the increased risk. The lack of difference in severe COVID-19 breakthrough illness risk between PWoH and PWH with higher CD4 counts may be due to engagement in medical care, different healthcare seeking behaviors and reduced comorbidities among the PWH included compared to PWoH. The increased risk of severe COVID-19 breakthrough illness for PWH with moderate (CD4 200-349 cells/mm^3^) immune suppression suggests they should be included with those who have advanced or untreated HIV in recommendations for additional primary series vaccination doses, second booster doses, and counseled on risk-reduction strategies.

Sex, age, comorbidities, and additional vaccine doses impact the risk of severe COVID-19 breakthrough illness.^10,31-33^ Among both PWoH and PWH, increasing age was the most significant risk factor for severe COVID-19 breakthrough illness in our study. Female PWoH had reduced risk, which has been previously documented;^34-36^ however, female PWH had increased severe risk. It is known that males and females have distinct immune system responses with females often demonstrating increased immune competence and less inflammatory immune responses, possibly contributing to their reduced risk for severe COVID-19 breakthrough illness; however, immune dysfunction with HIV may alter this effect and requires further investigation.^34,37,38^ Despite recommendations for additional COVID-19 vaccine doses being based in-part on CD4 count,^25^ we identified that the proportion of PWH who received additional doses varied little by CD4 count and was likely driven by clinical decision making and patient preference.

Among PWoH, several comorbidities have been associated with increased severe COVID-19 breakthrough illness risk.^32,33,39^ Our findings suggest an increased risk with HTN, ESRD, and SOT. Comorbidities were prevalent among those who experienced a severe COVID-19 breakthrough illness with 90% having a diagnosis of obesity, DM, HTN or ESRD. A lower proportion of PWH had at least one comorbidity than PWoH (82% vs 94%), yet their severe COVID-19 breakthrough rates remained the same as PWoH. Moderate to severe immune suppression from HIV itself is an important comorbidity that increases severe COVID-19 breakthrough risk; additional comorbidities and a recent cancer diagnosis increased severe COVID-19 breakthrough risk in PWH.

### Limitations

Our findings may not be generalizable to all PWH, as our study population had a greater proportion of males (89%) than found in the US population of PWH, and those with higher barriers to accessing healthcare (who may be at greater risk for COVID-19) were less likely to be included in our study population. Other outcome data, including mechanical ventilation and death (particularly if death occurred out of hospital) may be under-ascertained. The discharge diagnosis ranking was not consistent as one cohort was only able to provide the primary diagnosis with the remainder unranked; however, a sensitivity analyses demonstrated no significant differences in results following exclusion of this cohort. All discharge diagnoses were reviewed by clinicians to increase specificity in our classification of COVID-19 hospitalization, but discharge coding can be influenced by many factors including reimbursement practices. Similarly, our matching schema was not consistent, but the distributions of matching factors indicate that our sample of PWH and PWoH were comparable; we included the matching factors in multivariable analyses to address residual confounding.

### Conclusions

It was uncommon for COVID-19 breakthrough illness to progress to severe illness in our population of PWH and PWoH; however, PWH with moderate immune suppression (200-349 cells/mm^3^) had an increased severe COVID-19 breakthrough illness risk (compared to PWoH) and may benefit from being included in the CDC’s recommendation for those with “advanced and untreated HIV” to receive an additional dose in the primary COVID-19 vaccination series and second booster vaccination. Clinicians should continue to promote risk-reduction measures among PWH. The increased risk of severe COVID-19 breakthrough illness in PWH with moderate and severe immune suppression merits ongoing surveillance to inform vaccine recommendations as the pandemic persists, immunity to primary vaccine series and booster doses wane, and new variants emerge.

## Data Availability

Complete data for this study cannot be publicly shared because of legal and ethical restrictions.

## SUPPLEMENT

**Supplement Table 1:** Definitions of in care and criteria used to identify people with (PWH) and without HIV (PWoH)

| Cohort | Description | “In care” or “in cohort” | PWH |
| --- | --- | --- | --- |
| Kaiser Permanente Mid-Atlantic States (KPMAS) | Integrated Health system | In Care – KPMAS membership for ≥1 month in 2020 | HIV registry* |
| Kaiser Permanente Northern California (KPNC) | Integrated Health system | In Care – KPNC membership for ≥1 month between 3/1/20 and 12/31/20 | HIV registry* |
| University of North Carolina Chapel Hill (UNC) | Medical center cohort | ≥1 encounter with UNC Health in 2019 and alive as of 03/01/2020 | HIV diagnosis (by ICD diagnosis code) |
| Veterans Aging Cohort Study (VACS) | National cohort of all PWH and 1:2 demographically-matched PWoH in care in the Veterans Health Administration (VA) system | Enrolled in VACS from 1996-2017 and alive in 2020 | HIV diagnosis (presence of 1 inpatient or 2 outpatient ICD codes for HIV) |

The Kaiser Permanente Mid-Atlantic States and Northern California HIV Registries are databases of members diagnosed with HIV since 1998 and 1980, respectively. Primary sources used to identify HIV patients are HIV-specific laboratory tests, antiretroviral therapy, hospital-based HIV diagnosis, and diagnosis.

**Supplement Table 2:**
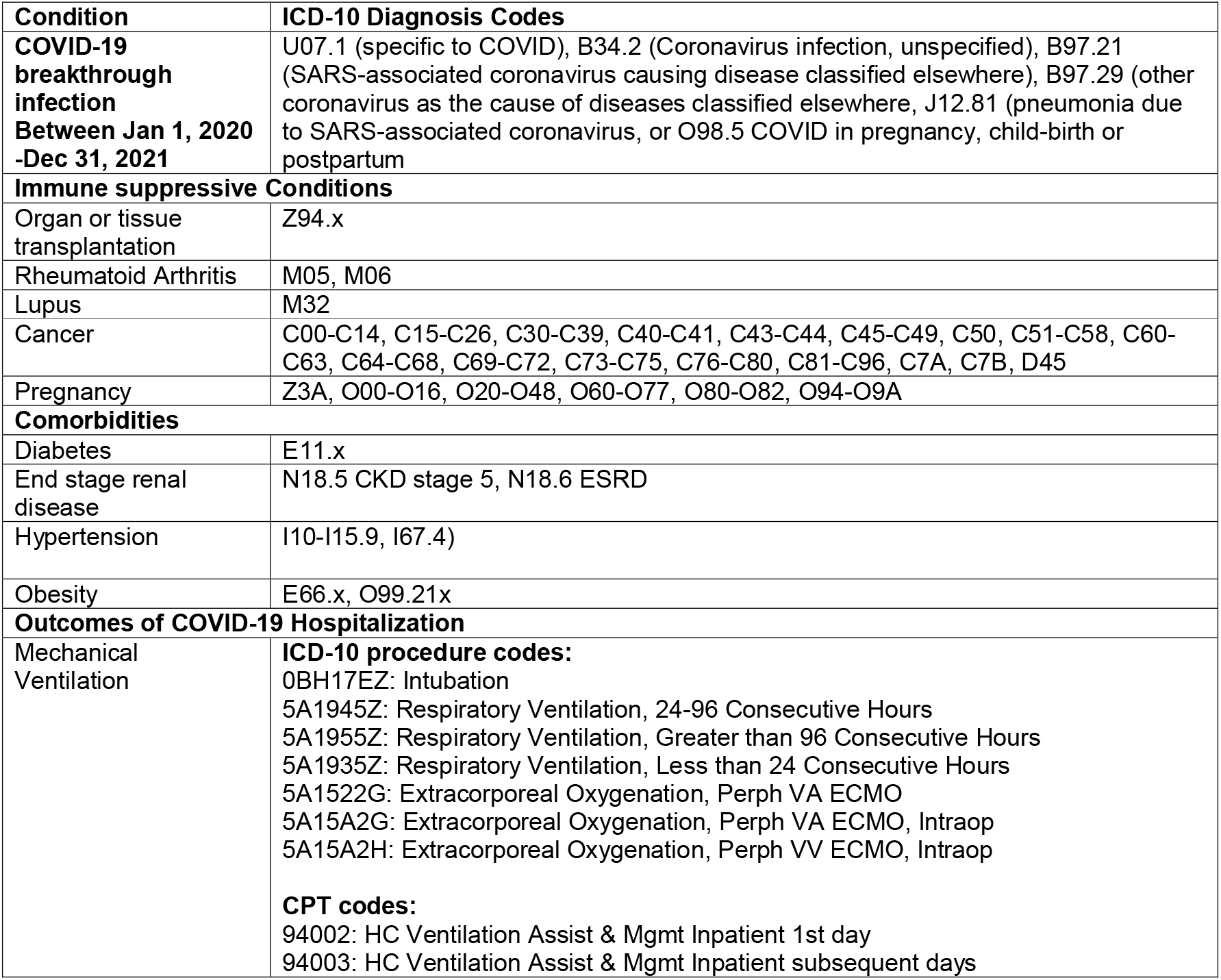
Diagnosis and Procedure codes used to ascertain comorbidities and outcomes

| **Condition** | **ICD-10 Diagnosis Codes** |
| --- | --- |
| **COVID-19 breakthrough infection Between Jan 1, 2020 -Dec 31, 2021** | U07.1 (specific to COVID), B34.2 (Coronavirus infection, unspecified), B97.21 (SARS-associated coronavirus causing disease classified elsewhere), B97.29 (other coronavirus as the cause of diseases classified elsewhere, J12.81 (pneumonia due to SARS-associated coronavirus, or O98.5 COVID in pregnancy, child-birth or postpartum |
| **Immune suppressive Conditions** |  |
| Organ or tissue transplantation | Z94.x |
| Rheumatoid Arthritis | M05, M06 |
| Lupus | M32 |
| Cancer | C00-C14, C15-C26, C30-C39, C40-C41, C43-C44, C45-C49, C50, C51-C58, C60-C63, C64-C68, C69-C72, C73-C75, C76-C80, C81-C96, C7A, C7B, D45 |
| Pregnancy | Z3A, O00-O16, O20-O48, O60-O77, O80-O82, O94-O9A |
| **Comorbidities** |  |
| Diabetes | E11.x |
| End stage renal disease | N18.5 CKD stage 5, N18.6 ESRD |
| Hypertension | I10-I15.9, I67.4) |
| Obesity | E66.x, O99.21x |
| **Outcomes of COVID-19 Hospitalization** |  |
| Mechanical Ventilation | **ICD-10 procedure codes:**<br>0BH17EZ: Intubation<br>5A1945Z: Respiratory Ventilation, 24-96 Consecutive Hours<br>5A1955Z: Respiratory Ventilation, Greater than 96 Consecutive Hours<br>5A1935Z: Respiratory Ventilation, Less than 24 Consecutive Hours<br>5A1522G: Extracorporeal Oxygenation, Perph VA ECMO<br>5A15A2G: Extracorporeal Oxygenation, Perph VA ECMO, Intraop<br>5A15A2H: Extracorporeal Oxygenation, Perph VV ECMO, Intraop<br>**CPT codes:**<br>94002: HC Ventilation Assist & Mgmt Inpatient 1st day<br>94003: HC Ventilation Assist & Mgmt Inpatient subsequent days |

**Supplement Table 3:**
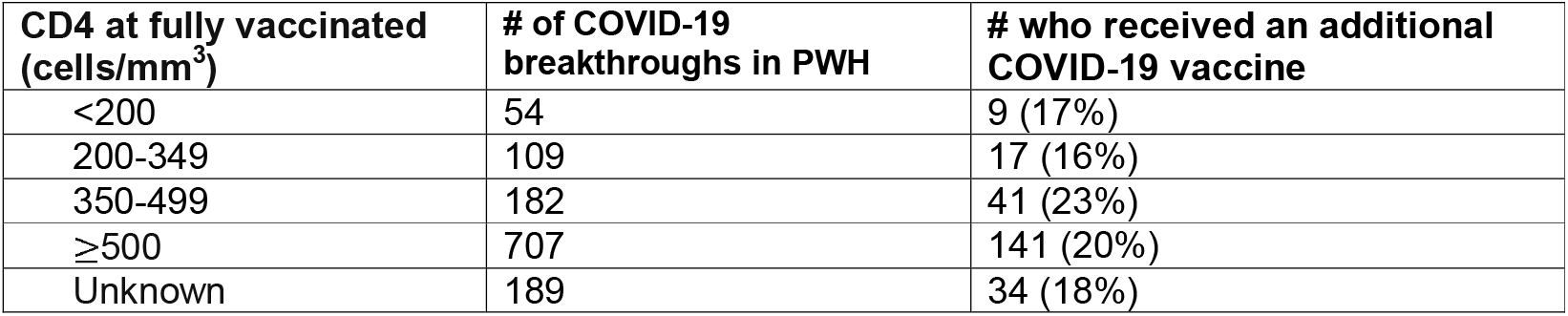
Proportion of PWH who received additional vaccine doses by CD4 count at fully vaccinated (N=1,241).

| **CD4 at fully vaccinated**<br>**(cells/mm<sup>3</sup>)** | **# of COVID-19**<br>**breakthroughs in PWH** | **# who received an additional**<br>**COVID-19 vaccine** |
| --- | --- | --- |
| <200 | 54 | 9 (17%) |
| 200-349 | 109 | 17 (16%) |
| 350-499 | 182 | 41 (23%) |
| ≥500 | 707 | 141 (20%) |
| Unknown | 189 | 34 (18%) |

**Supplement Table 4:** Characteristics of SARS-CoV-2 breakthrough infection, both severe and non-severe by HIV status N=3,649

|  | PWoH N=2,408 |  | PWH N=1,241 |  |
| --- | --- | --- | --- | --- |
| Characteristic | No Severe Breakthrough<br>N = 2,238 | Severe Breakthrough<br>N = 170 | No Severe Breakthrough<br>N = 1,162 | Severe Breakthrough<br>N = 79 |
| **Age (years)** |  |  |  |  |
| <55 | 907 (41%) | 10 (6%) | 534 (46%) | 16 (20%) |
| 55-64 | 741 (33%) | 56 (33%) | 333 (28%) | 26 (33%) |
| 65-74 | 486 (21%) | 71 (42%) | 240 (21%) | 24 (30%) |
| 75+ | 122 (5%) | 33 (19%) | 55 (5%) | 13 (16%) |
| **Sex** |  |  |  |  |
| Male | 1944 (87%) | 164 (97%) | 1,065 (91%) | 71 (90%) |
| Female | 294 (13%) | 6 (4%) | 97 (8%) | 8 (10%) |
| **Ethnicity and Race** |  |  |  |  |
| Non-Hispanic White | 694 (31%) | 50 (29%) | 397 (34%) | 24 (30%) |
| Non-Hispanic Black/<br>African American | 1,060 (47%) | 91 (54%) | 515 (44%) | 40 (51%) |
| Hispanic | 342 (15%) | 24 (14%) | 173 (15%) | 10 (13%) |
| Other | 127 (6%) | 4 (3%) | 66 (6%) | 5 (6%) |
| Unknown | 15 (1%) | 1 (1%) | 11 (1%) | 0 (0.0%) |
| **Month of breakthrough** |  |  |  |  |
| Jan-June | 231 (10%) | 16 (9%) | 129 (11%) | 5 (6%) |
| July-Oct | 1,024 (46%) | 108 (64%) | 585 (50%) | 54 (68%) |
| Nov-Dec | 983 (44%) | 46 (27%) | 448 (39%) | 20 (25%) |
| **Vaccine series** |  |  |  |  |
| Pfizer | 1,103 (49%) | 91 (54%) | 559 (48%) | 46 (58%) |
| Pfizer + 3rd dose | 187 (8%) | 7 (4%) | 160 (14%) | 5 (6%) |
| Moderna | 629 (28%) | 47 (28%) | 285 (25%) | 16 (20%) |
| Moderna + 3rd dose | 90 (4%) | 5 (3%) | 67 (6%) | 1 (1%) |
| J&J | 215 (10%) | 19 (11%) | 82 (7%) | 11 (14%) |
| J&J + 2<sup>nd</sup> dose | 14 (1%) | 1 (1%) | 9 (1%) | 0 (0%) |
| **COVID prior to full vaccination date** | 283 (13%) | 9 (5%) | 170 (15%) | 6 (8%) |
| **Comorbidities before fully vaccinated** |  |  |  |  |
| Obese BMI ($\geq 30$ kg/m<sup>2</sup>) | 1,132 (54%) | 86 (53%) | 402 (36%) | 32 (43%) |
| Diabetes | 718 (32%) | 99 (58%) | 276 (24%) | 32 (41%) |
| Hypertension | 1,287 (58%) | 156 (92%) | 577 (50%) | 56 (71%) |
| End-stage renal disease | 41 (2%) | 22 (13%) | 34 (3%) | 4 (5%) |
| Organ or tissue transplantation | 27 (1%) | 16 (9%) | 17 (1%) | 1 (1%) |
| Rheumatoid arthritis | 38 (2%) | 6 (4%) | 13 (1%) | 1 (1%) |
| Lupus | 8 (0%) | 2 (1%) | 3 (0%) | 0 (0%) |
| Characteristic | No Severe Breakthrough<br>N = 2,238 | Severe Breakthrough<br>N = 170 | No Severe Breakthrough<br>N = 1,162 | Severe Breakthrough<br>N = 79 |
| Cancer diagnosis | 157 (7%) | 30 (18%) | 111 (10%) | 18 (23%) |
| Pregnancy | 4 (0%) | 0 (0%) | 3 (0%) | 0 (0%) |

Age was categorized into a 4-level group to assess a dose response relationship among older people (i.e., 55+) that are known to be at greater risk.

**Supplement Figure 1:**
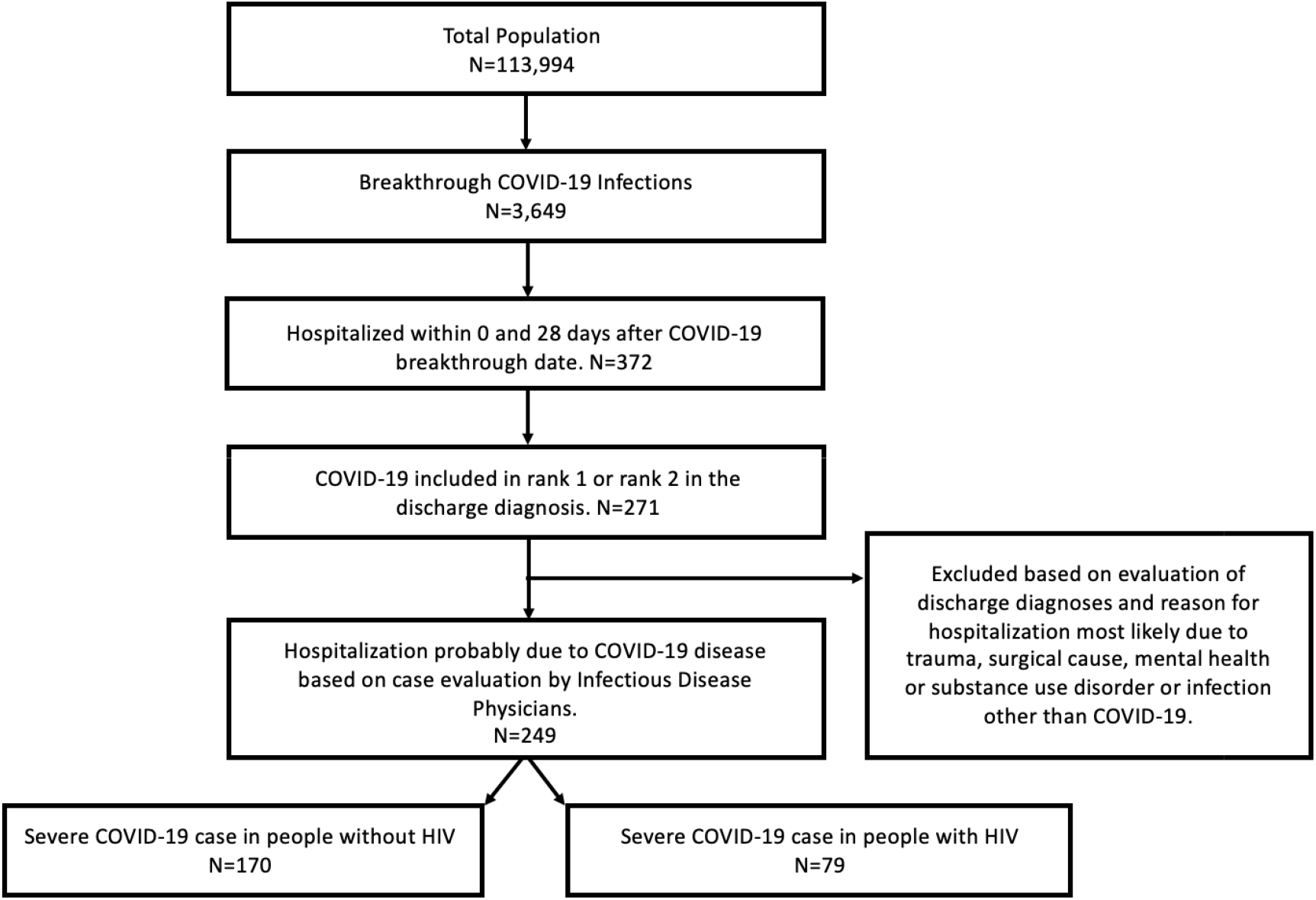
Identification of severe COVID-19 breakthrough infections

**Supplement Table 5:** Severe COVID-19 breakthrough illness Incidence rates and 95% confidence intervals, by month and HIV status

|  |  | # of severe COVID-19 breakthroughs | # of person-years | IR (95% CI) per 100 person years |
| --- | --- | --- | --- | --- |
| **Total** |  |  |  |  |
|  | Overall | 249 | 191.3 | 137.5 (117.6, 159.8) |
|  | PWH | 79 | 67.7 | 116.7 (92.4, 145.4) |
|  | PWoH | 170 | 123.6 | 130.2 (114.5, 147.4) |
| **Moderna** |  |  |  |  |
|  | Overall | 69 | 61.9 | 111.5 (86.8, 141.2) |
|  | PWH | 17 | 21.0 | 81.1 (47.3, 129.9) |
|  | PWoH | 52 | 40.9 | 127.1 (94.9, 166.7) |
| **Pfizer** |  |  |  |  |
|  | Overall | 149 | 110.3 | 135.1 (114.3, 158.7) |
|  | PWH | 51 | 41.1 | 124.1 (92.4, 163.2) |
|  | PWoH | 98 | 69.2 | 141.7 (115.0, 172.7) |
| **J&J** |  |  |  |  |
|  | Overall | 31 | 19.2 | 161.5 (109.7, 229.2) |
|  | PWH | 11 | 5.7 | 194.4 (97.0, 347.8) |
|  | PWoH | 20 | 13.5 | 147.8 (90.2, 228.2) |

| **By calendar month** |  |  |  |  |
| --- | --- | --- | --- | --- |
| Jan | Overall | 0 | 0.02 | 0.0 (0.0, 19248.5) |
|  | PWH | 0 | 0 | n/a |
|  | PWoH | 0 | 0.02 | 0.0 (0.0, 19248.1) |
| Feb | Overall | 0 | 0.35 | 0.00 (0.00, 1052.6) |
|  | PWH | 0 | 0.14 | 0.0 (0.0, 2641.9) |
|  | PWoH | 0 | 0.21 | 0.0 (0.0, 1749.8) |
| Mar | Overall | 2 | 2.56 | 78.2 (9.5, 282.5) |
|  | PWH | 0 | 0.90 | 0.0 (0.0, 409.5) |
|  | PWoH | 2 | 1.66 | 120.7 (14.6, 436.2) |
| Apr | Overall | 6 | 6.06 | 99.0 (36.3, 215.5) |
|  | PWH | 1 | 2.13 | 47.0 (1.2, 261.6) |
|  | PWoH | 5 | 3.93 | 127.2 (41.3, 296.8) |
| May | Overall | 8 | 9.07 | 88.2 (38.1, 173.8) |
|  | PWH | 3 | 3.13 | 95.8 (19.8, 272.9) |
|  | PWoH | 5 | 5.94 | 84.2 (27.3, 196.5) |
| June | Overall | 3 | 7.33 | 41.0 (8.4, 119.7) |
|  | PWH | 1 | 2.52 | 39.7 (1.0, 221.2) |
|  | PWoH | 2 | 4.81 | 41.6 (5.0, 150.3) |
| Jul | Overall | 16 | 13.25 | 120.7 (69.0, 196.1) |
|  | PWH | 4 | 5.60 | 71.4 (19.5, 182.7) |
|  | PWoH | 12 | 7.65 | 156.9 (81.1, 274.1) |
| Aug | Overall | 69 | 44.68 | 154.4 (120.2, 195.4) |
|  | PWH | 25 | 19.2 | 130.2 (84.3, 192.2) |
|  | PWoH | 44 | 25.5 | 172.7 (125.5, 231.8) |
| Sept | Overall | 57 | 42.40 | 134.5 (101.8, 174.2) |
|  | PWH | 18 | 13.9 | 129.9 (77.0, 205.3) |
|  | PWoH | 39 | 28.5 | 136.7 (97.2, 186.8) |
|  | Overall | 20 | 29.25 | 68.4 (41.8, 105.6) |
| Oct | PWH | 7 | 9.5 | 74.1 (29.8, 152.7) |
|  | PWoH | 13 | 19.8 | 65.7 (35.0, 112.3) |
| Nov | Overall | 24 | 19.15 | 125.3 (80.3, 186.5) |
|  | PWH | 7 | 6.0 | 117.3 (47.2, 241.8) |
|  | PWoH | 17 | 13.2 | 128.9 (75.1, 206.4) |
| Dec | Overall | 44 | 36.67 | 120.0 (87.2, 161.1) |
|  | PWH | 13 | 11.4 | 114.4 (60.9, 195.7) |
|  | PWoH | 31 | 25.3 | 122.5 (83.2, 173.8) |

**Supplement Figure 2:**
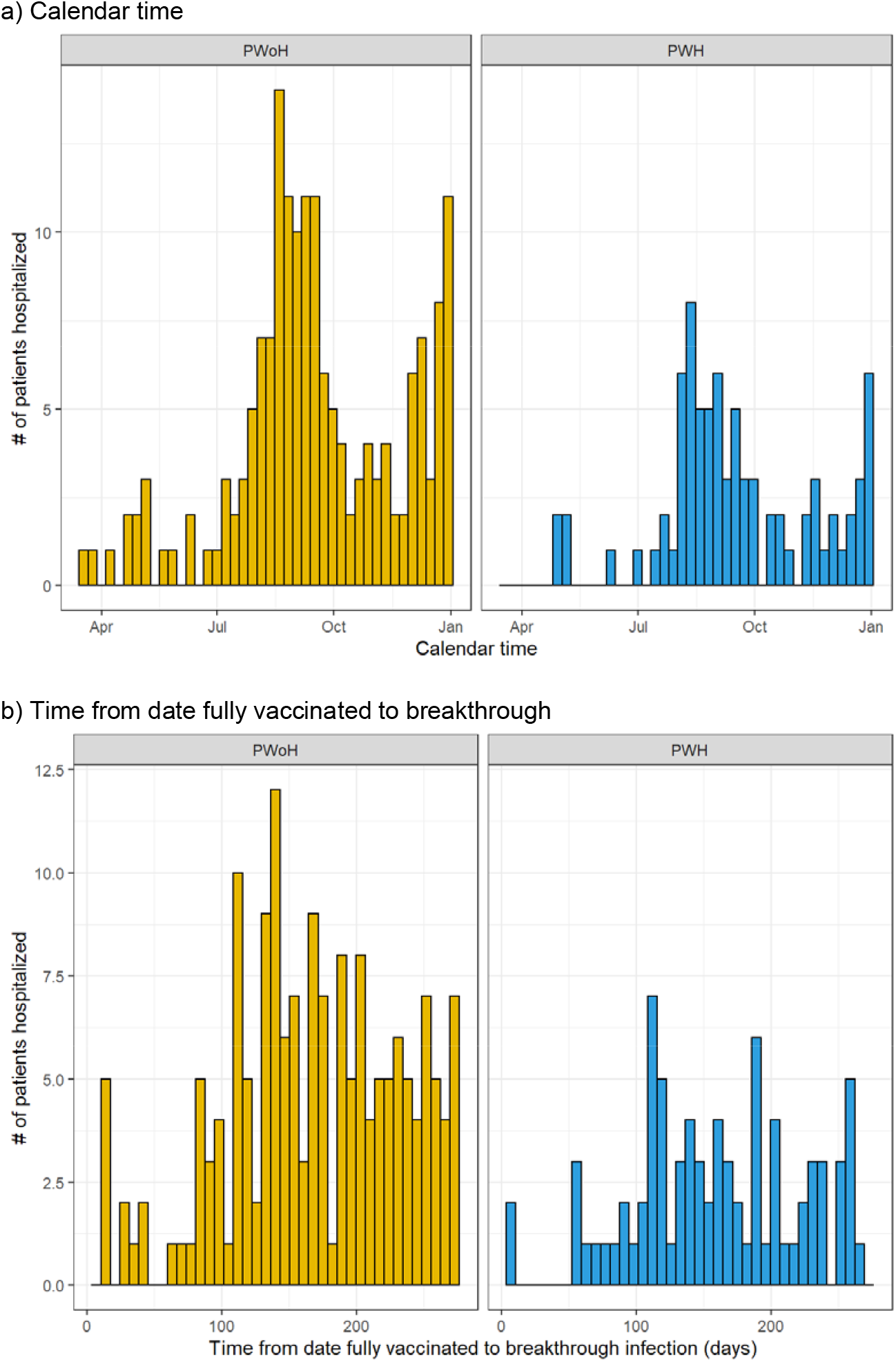
Histograms of the date of severe SARS-CoV-2 breakthrough infection, by HIV status

**Supplemental Figure 3:**
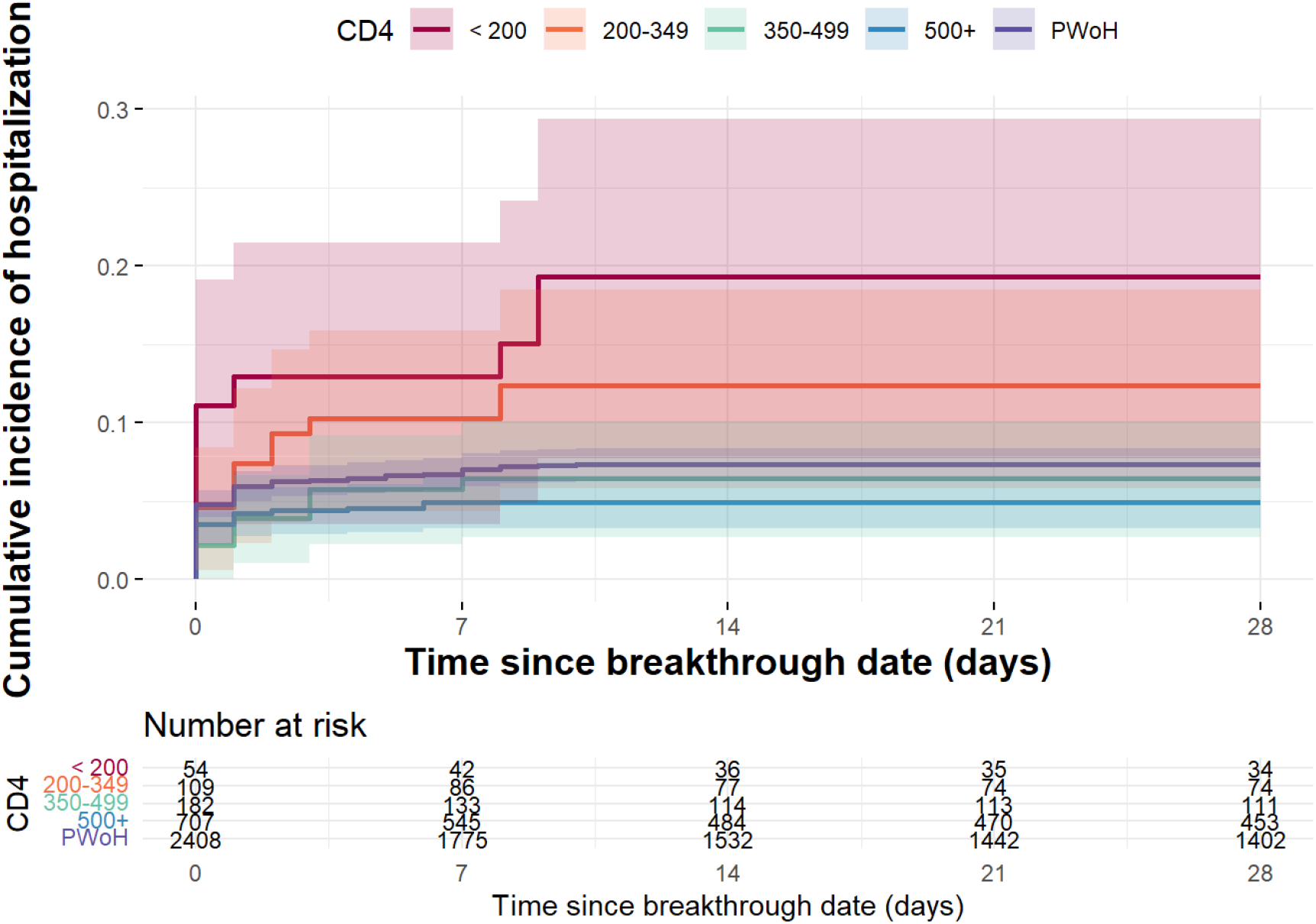
Cumulative incidence of severe COVID-19 breakthrough illness (and 95% confidence intervals represented by the shading) stratified by CD4 count and HIV status

**Supplement Table 6:** Characteristics of participants with breakthrough COVID-19 illness requiring hospitalization, mechanical ventilation or in those who died

| Characteristic | Hospitalized<br>(Severe COVID-19)<br>**N = 249** | Mechanical<br>ventilation<br>**N = 24** | Death<br>**N = 20** |
| --- | --- | --- | --- |
| **HIV Status** |  |  |  |
| PWoH | 170 (68%) | 16 (67%) | 14 (70%) |
| PWH | 79 (32%) | 8 (33%) | 6 (30%) |
| **Age (years)** |  |  |  |
| <55 | 19 (8%) | 3 (13%) | 2 (10%) |
| 55-64 | 82 (33%) | 9 (38%) | 4 (20%) |
| 65-74 | 95 (38%) | 6 (25%) | 5 (25%) |
| 75+ | 46 (19%) | 6 (25%) | 9 (45%) |
| **Sex** |  |  |  |
| Male | 235 (94%) | 20 (83%) | 18 (90%) |
| Female | 14 (6%) | 4 (17%) | 2 (10%) |
| **Ethnicity and Race** |  |  |  |
| Non-Hispanic White | 74 (30%) | 2 (8%) | 5 (25%) |
| Non-Hispanic Black/African American | 131 (53%) | 18 (75%) | 11 (55%) |
| Hispanic | 34 (14%) | 4 (17%) | 4 (20%) |
| Other | 9 (4%) | 0 (0%) | 0 (0%) |
| Unknown | 1 (0%) | 0 (0%) | 0 (0%) |
| **Month of breakthrough** |  |  |  |
| Jan-June | 21 (8%) | 3 (13%) | 3 (15%) |
| July-Oct | 162 (65%) | 16 (67%) | 13 (65%) |
| Nov-Dec | 66 (27%) | 5 (21%) | 4 (20%) |
| **COVID prior to vaccination** | 15 (6%) | 0 (0%) | 0 (0%) |
| **Vaccine Series** |  |  |  |
| Primary Series Only | 230 (92%) | 21 (88%) | 18 (90%) |

|  | Hospitalized<br>(Severe COVID-19) | Mechanical<br>ventilation | Death |
| --- | --- | --- | --- |
| **Characteristic** | **N = 249** | **N = 24** | **N = 20** |
| Primary Series + additional dose | 19 (8%) | 3 (13%) | 2 (10%) |
| **Obese BMI ($\geq 30$ kg/m<sup>2</sup>)** | 118 (50%) | 20 (87%) | 13 (68%) |
| Unknown | 13 | 1 | 1 |
| **Diabetes** | 131 (53%) | 15 (63%) | 13 (65%) |
| **Hypertension** | 212 (85%) | 23 (96%) | 19 (95%) |
| **End-stage renal disease** | 26 (10%) | 5 (21%) | 6 (30%) |
| **Organ or tissue transplantation** | 17 (7%) | 5 (21%) | 4 (20%) |
| **Rheumatoid arthritis** | 7 (3%) | 3 (13%) | 2 (10%) |
| **Lupus** | 2 (1%) | 0 (0%) | 0 (0%) |
| **Cancer diagnosis** | 48 (19%) | 5 (21%) | 5 (25%) |
| **CD4 at ART initiation** | 399 (212,546) | 376 (324,417) | 398 (354,855) |
| Unknown | 210 | 20 | 15 |
| **AIDS before fully vaccinated** | 29 (37%) | 3 (38%) | 3 (50%) |
| Unknown | 170 | 16 | 14 |
| **CD4 at fully vaccinated** | 498 (305,746) | 352 (172,542) | 352 (291,423) |
| Unknown | 181 | 18 | 16 |
| **Suppressed HIV RNA at fully vaccinated** | 59 (87%) | 5 (83%) | 4 (100%) |
| Unknown | 181 | 18 | 16 |

n(%); median (IQR)

**Supplementa Figure 4:**
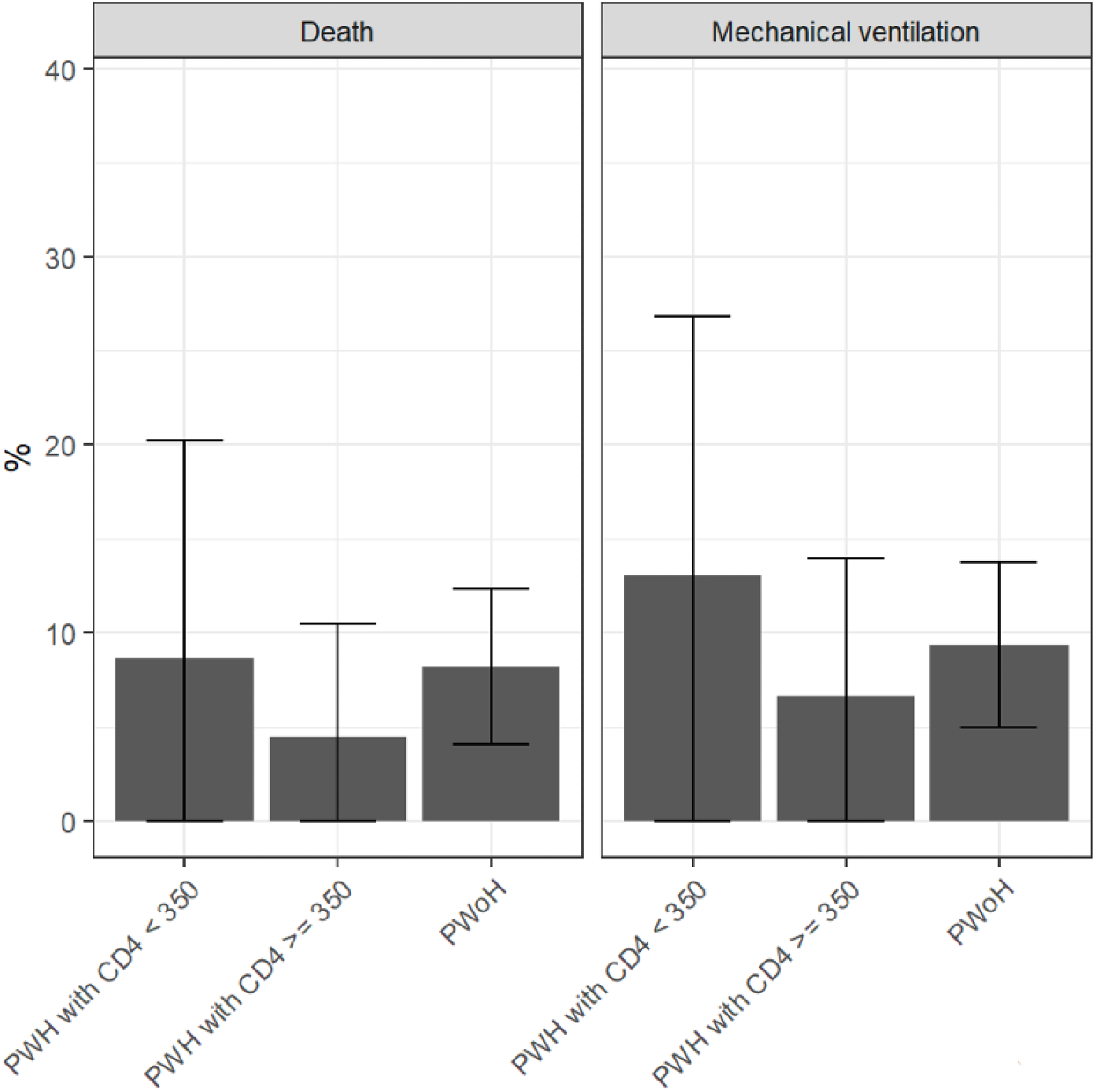
Outcomes of breakthrough illness among those hospitalized after fully vaccinated to Dec 31, 2021, by HIV status (n=170 PWoH) and CD4 count (n=23 <350 cells/mm^3^ and n=45 ≥350 cells/mm^3^) among PWH

## REFERENCES

1. Polack FP, Thomas SJ, Kitchin N, et al. Safety and Efficacy of the BNT162b2 mRNA Covid-19 Vaccine. N Engl J Med. Dec 31 2020;383(383):2603–2615. doi:10.1056/NEJMoa2034577

2. Baden LR, El Sahly HM, Essink B, et al. Efficacy and Safety of the mRNA-1273 SARS-CoV-2 Vaccine. N Engl J Med. Feb 4 2021;384(384):403–416. doi:10.1056/NEJMoa2035389

3. Sadoff J, Gray G, Vandebosch A, et al. Safety and Efficacy of Single-Dose Ad26.COV2.S Vaccine against Covid-19. N Engl J Med. Jun 10 2021;384(384):2187–2201. doi:10.1056/NEJMoa2101544

4. Coburn SB, Humes E, Lang R, et al. COVID-19 infections post-vaccination by HIV status in the United States. medRxiv. Dec 5 2021;doi:10.1101/2021.12.02.21267182

5. Sun J, Zheng Q, Madhira V, et al. Association Between Immune Dysfunction and COVID-19 Breakthrough Infection After SARS-CoV-2 Vaccination in the US. JAMA Intern Med. Dec 28 2021;doi:10.1001/jamainternmed.2021.7024

6. Kim AHJ, Nakamura MC. COVID-19 Breakthrough Infection Among Immunocompromised Persons. JAMA Intern Med. Dec 28 2021;doi:10.1001/jamainternmed.2021.7033

7. Yamamoto S, Maeda K, Matsuda K, et al. COVID-19 breakthrough infection and post-vaccination neutralizing antibody among healthcare workers in a referral hospital in Tokyo: a case-control matching study. Clin Infect Dis. Dec 24 2021;doi:10.1093/cid/ciab1048

8. Team CC-VBCI. COVID-19 Vaccine Breakthrough Infections Reported to CDC - United States, January 1-April 30, 2021. MMWR Morb Mortal Wkly Rep. May 28 2021;70(70):792–793. doi:10.15585/mmwr.mm7021e3

9. Haidar G, Agha M, Bilderback A, et al. Prospective evaluation of COVID-19 vaccine responses across a broad spectrum of immunocompromising conditions: the COVICS study. Clin Infect Dis. Feb 18 2022;doi:10.1093/cid/ciac103

10. Butt AA, Yan P, Shaikh OS, Mayr FB. Outcomes among patients with breakthrough SARS-CoV-2 infection after vaccination in a high-risk national population. EClinicalMedicine. Oct 2021;40:101117. doi:10.1016/j.eclinm.2021.101117

11. Lin DY, Gu Y, Wheeler B, et al. Effectiveness of Covid-19 Vaccines over a 9-Month Period in North Carolina. N Engl J Med. Mar 10 2022;386(386):933–941. doi:10.1056/NEJMoa2117128

12. Collins LF, Moran CA, Oliver NT, et al. Clinical characteristics, comorbidities and outcomes among persons with HIV hospitalized with coronavirus disease 2019 in Atlanta, Georgia. AIDS. Oct 1 2020;34(34):1789–1794. doi:10.1097/QAD.0000000000002632

13. Shalev N, Scherer M, LaSota ED, et al. Clinical Characteristics and Outcomes in People Living With Human Immunodeficiency Virus Hospitalized for Coronavirus Disease 2019. Clin Infect Dis. Nov 19 2020;71(71):2294–2297. doi:10.1093/cid/ciaa635

14. Ceballos ME, Ross P, Lasso M, et al. Clinical characteristics and outcomes of people living with HIV hospitalized with COVID-19: a nationwide experience. Int J STD AIDS. Apr 2021;32(32):435–443. doi:10.1177/0956462420973106

15. Durstenfeld MS, Sun K, Ma Y, et al. Association of HIV infection with outcomes among adults hospitalized with COVID-19. AIDS. Mar 1 2022;36(36):391–398. doi:10.1097/QAD.0000000000003129

16. Bhaskaran K, Rentsch CT, MacKenna B, et al. HIV infection and COVID-19 death: a population-based cohort analysis of UK primary care data and linked national death registrations within the OpenSAFELY platform. Lancet HIV. Jan 2021;8(8):e24–e32. doi:10.1016/S2352-3018(20)30305-2

17. Tesoriero JM, Swain CE, Pierce JL, et al. COVID-19 Outcomes Among Persons Living With or Without Diagnosed HIV Infection in New York State. JAMA Netw Open. Feb 1 2021;4(4):e2037069. doi:10.1001/jamanetworkopen.2020.37069

18. Yang X, Sun J, Patel RC, et al. Associations between HIV infection and clinical spectrum of COVID-19: a population level analysis based on US National COVID Cohort Collaborative (N3C) data. Lancet HIV. Nov 2021;8(8):e690–e700. doi:10.1016/S2352-3018(21)00239-3

19. Dong Y, Li Z, Ding S, et al. HIV infection and risk of COVID-19 mortality: A meta-analysis. Medicine (Baltimore). Jul 2 2021;100(100):e26573. doi:10.1097/MD.0000000000026573

20. Cooper TJ, Woodward BL, Alom S, Harky A. Coronavirus disease 2019 (COVID-19) outcomes in HIV/AIDS patients: a systematic review. HIV Med. Oct 2020;21(21):567–577. doi:10.1111/hiv.12911

21. Hoffmann C, Casado JL, Harter G, et al. Immune deficiency is a risk factor for severe COVID-19 in people living with HIV. HIV Med. May 2021;22(22):372–378. doi:10.1111/hiv.13037

22. Lesko CR, Bengtson AM. HIV and COVID-19: Intersecting Epidemics With Many Unknowns. Am J Epidemiol. Jan 4 2021;190(190):10–16. doi:10.1093/aje/kwaa158

23. Hadi YB, Naqvi SFZ, Kupec JT, Sarwari AR. Characteristics and outcomes of COVID-19 in patients with HIV: a multicentre research network study. AIDS. Nov 1 2020;34(34):F3–F8. doi:10.1097/QAD.0000000000002666

24. Centers for Disease Control and Prevention. COVID-19 Vaccines for Moderately to Severely Immunocompromised People Updated Feb 17, 2022. Accessed March 14, 2021. https://www.cdc.gov/coronavirus/2019-ncov/vaccines/recommendations/immuno.html

25. Centers for Disease Control and Prevention (CDC). COVID-19 and HIV. Accessed March 31, 2022. https://www.cdc.gov/hiv/covid-19/index.html

26. Gange SJ, Kitahata MM, Saag MS, et al. Cohort profile: the North American AIDS Cohort Collaboration on Research and Design (NA-ACCORD). Int J Epidemiol. Apr 2007;36(36):294–301. doi:10.1093/ije/dyl286

27. When You’ve Been Fully Vaccinated | CDC. https://www.cdc.gov/coronavirus/2019-ncov/vaccines/fully-vaccinated.html. Accessed January 4, 2022.

28. Fultz SL, Skanderson M, Mole LA, et al. Development and verification of a “virtual” cohort using the National VA Health Information System. Med Care. Aug 2006;44(8 Suppl 2):S25–30. doi:10.1097/01.mlr.0000223670.00890.74

29. Investigative Criteria for Suspected Cases of SARS-CoV-2 Reinfection (ICR) | CDC. https://www.cdc.gov/coronavirus/2019-ncov/php/invest-criteria.html. Accessed January 4, 2022.

30. Centers for Disease Control and Prevention. AIDS-Defining Conditions. MMWR Recommendations and Reports. https://www.cdc.gov/mmwr/preview/mmwrhtml/rr5710a2.htmPublished December 5, 2008. Accessed January 4, 2021.

31. Botton J, Semenzato L, Jabagi MJ, et al. Effectiveness of Ad26.COV2.S Vaccine vs BNT162b2 Vaccine for COVID-19 Hospitalizations. JAMA Netw Open. Mar 1 2022;5(5):e220868. doi:10.1001/jamanetworkopen.2022.0868

32. Wright BJ, Tideman S, Diaz GA, French T, Parsons GT, Robicsek A. Comparative vaccine effectiveness against severe COVID-19 over time in US hospital administrative data: a case-control study. Lancet Respir Med. Feb 25 2022;doi:10.1016/S2213-2600(22)00042-X

33. Fabiani M, Puopolo M, Morciano C, et al. Effectiveness of mRNA vaccines and waning of protection against SARS-CoV-2 infection and severe covid-19 during predominant circulation of the delta variant in Italy: retrospective cohort study. BMJ. Feb 10 2022;376:e069052. doi:10.1136/bmj-2021-069052

34. Pradhan A, Olsson PE. Sex differences in severity and mortality from COVID-19: are males more vulnerable? Biol Sex Differ. Sep 18 2020;11(11):53. doi:10.1186/s13293-020-00330-7

35. Vahidy FS, Pan AP, Ahnstedt H, et al. Sex differences in susceptibility, severity, and outcomes of coronavirus disease 2019: Cross-sectional analysis from a diverse US metropolitan area. PLoS One. 2021;16(16):e0245556. doi:10.1371/journal.pone.0245556

36. Liu C, Lee J, Ta C, et al. A Retrospective Analysis of COVID-19 mRNA Vaccine Breakthrough Infections - Risk Factors and Vaccine Effectiveness. medRxiv. Oct 7 2021;doi:10.1101/2021.10.05.21264583

37. Strandberg TE, Pentti J, Kivimaki M. Sex Difference in Serious Infections: Not Only COVID-19. Epidemiology. Nov 1 2021;32(32):e26–e27. doi:10.1097/EDE.0000000000001408

38. Qi S, Ngwa C, Morales Scheihing DA, et al. Sex differences in the immune response to acute COVID-19 respiratory tract infection. Biol Sex Differ. Dec 20 2021;12(12):66. doi:10.1186/s13293-021-00410-2

39. Butt AA, Yan P, Shaikh OS, Mayr FB, Omer SB. Rate and Risk Factors for Severe/Critical Disease Among Fully Vaccinated Persons with Breakthrough SARS-CoV-2 Infection in a High-risk National Population. Clin Infect Dis. Dec 10 2021;doi:10.1093/cid/ciab1023

